# Investigating the Causal Impact of COVID-19 on Acute Myocardial Infarction: A Multi-ancestry Mendelian Randomization Study

**DOI:** 10.64898/2026.10.07.26364920

**Authors:** Katherine Zhou, Ying Ma

## Abstract

Observational studies suggest an association between COVID-19 and acute myocardial infarction (AMI), however, whether this relationship is causal remains unclear. We conducted a multi-ancestry Mendelian randomization (MR) study across European, East Asian, South Asian, and African populations, evaluating three COVID-19 phenotypes: very severe COVID-19, hospitalized COVID-19, and general infection association with AMI. GWAS summary statistics were obtained from the COVID-19 Host Genetics Initiative Round 7 and UK Biobank/Biobank Japan for AMI. Inverse-variance weighted (IVW) MR was the primary method, with fixed-or random-effects models selected according to heterogeneity test. MR-Egger, weighted median and weighted mode were conducted as sensitivity analyses. In European ancestry, genetic liability to very severe COVID-19 and hospitalized COVID-19 was slightly inversely associated with AMI risk, but without statistical significances across sensitivity analyses. Hospitalized COVID-19 showed a suggestive but non-significant inverse association in South Asian ancestry. No significant associations were identified in East Asian or African ancestries. Our findings did not support a robust causal association between COVID-19 susceptibility or severity and AMI risk across ancestries, suggesting that the observed cardiovascular burden may be driven more by acute pathophysiological mechanisms than shared genetic architecture.

## 1 Introduction

Cardiovascular disease remains the leading cause of mortality worldwide and accounts for a substantial proportion of global morbidity and death (Di Cesare et al. 2024). Although coronavirus disease 2019 (COVID-19), caused by the severe acute respiratory syndrome coronavirus-2 (SARS-CoV-2), primarily manifests as a respiratory illness, it is increasingly recognized as a systemic disease that can affect the cardiovascular system, contributing to conditions such as myocarditis, heart failure, and acute myocardial infarction (AMI) (Krittanawong et al. 2020; Krishna 2024). Myocardial injury has been reported in 5%-38% of hospitalized patients with COVID-19 and is associated with substantially increased mortality (Bavishi et al. 2020). The clinical burden is particularly high among patients with concurrent COVID-19 and AMI, with mortality rates of 10%–76% compared with 4.6%– 7% among AMI patients without COVID-19 (Rus et al. 2024). Large observational studies further indicate that AMI occurs in up to 8.9% of hospitalized COVID-19 patients, making it one of the most common atherothrombotic complications of the disease (Nechita et al. 2024).

Importantly, the cardiovascular consequences of COVID-19 may extend beyond the acute phase of infection. Epidemiological studies have reported a persistently increased risk of AMI among COVID-19 survivors for months to years after the initial infection (Zuin et al. 2023). A meta-analysis involving more than 20 million individuals estimated that COVID-19 survivors had an approximately 93% higher risk of AMI compared with the general population (Zuin et al. 2023). Several biological mechanisms have been proposed to link COVID-19 with AMI, including direct viral toxicity, cytokine storm, endothelial cell damage with thrombo-inflammation, and oxygen supply-demand mismatch (Dou et al. 2020). Nevertheless, these observational associations do not imply causality, as they remain susceptible to residual confounding (in which unmeasured or inadequately controlled factors, such as pre-existing cardiovascular risk, may account for part of the observed association) and reverse causation (in which individuals with subclinical or established cardiovascular disease may have been more susceptible to severe COVID-19 in the first place), thereby limiting the ability to draw definitive causal conclusions from these data.

Growing evidence suggests that COVID-19 severity and outcomes also vary substantially across racial and ethnic groups. Studies from the World Heart Federation COVID-19 Study and other large cohorts have demonstrated marked disparities in COVID-19 severity and mortality (Prabhakaran et al. 2022). Compared with individuals of European ancestry, Asian, Black, and Hispanic populations experience approximately two-to four-fold higher mortality risks during COVID-19 hospitalization (Prabhakaran et al. 2022). In a multicenter cohort of 613 patients admitted to six London hospitals, Asian and Afro-Caribbean patients demonstrated higher all-cause mortality and distinct cardiovascular complications, including myocardial injury, relative to European patients (Papageorgiou et al. 2020). These observed disparities may reflect differences in social determinants of health, comorbidity burden, healthcare access, environmental exposures, and biological factors. In addition, differences in allele frequencies, linkage disequilibrium patterns, and genetic architecture across ancestry groups may influence the validity and transportability of genetic analyses. These considerations motivate ancestry-stratified investigations of the relationship between genetic liability to COVID-19 and cardiovascular outcomes.

Traditional observational studies examining the association between COVID-19 and cardiovascular outcomes are not causal in nature, as they are inherently vulnerable to reverse causality and residual confounding factors (Wang et al. 2022). Mendelian randomization (MR) is an analytical method that uses naturally inherited genetic variants as instrumental variables to estimate the causal effect of an exposure on an outcome, based on the fact that genetic variants are randomly assigned at conception and remain fixed throughout a person’s lifetime (Sekula et al. 2016; Sanderson et al. 2022). The random allocation of genetic variants in MR resembles the logic of randomized controlled trials and helps reduce confounding and reverse causation because genetic variants are determined before disease development or environmental exposures (Burgess et al. 2019). To ensure the validity of the causal conclusion from MR, each selected instrumental variable needs to satisfy three MR modeling assumptions: (1) genetic variants are robustly associated with exposure (relevance); (2) they are independent of confounders (independence); and (3) they affect the outcome only through the exposure, without alternative biological pathways (exclusion restriction) (Sekula et al. 2016; Sanderson et al. 2022). Large-scale genome-wide association studies (GWASs) on COVID-19 performed in the recent years have identified many SNPs associated with COVID-19 across different ancestries, making it feasible to choose appropriate SNPs to serve as valid instruments for COVID-19 (COVID-19 Host Genetics Initiative. 2022).

Several MR methods exist, each with distinct assumptions and tolerances for pleiotropy. The inverse-variance weighted (IVW) method serves as the primary estimator and yields the most precise causal estimate when pleiotropy is absent, pooling the ratio estimates from each genetic variant weighted by their precision (Bowden et al. 2016). The MR-Egger method relaxes the exclusion restriction by allowing a non-zero intercept, which provides a test for directional pleiotropy and a bias-corrected causal estimate under the InSIDE assumption (Dudbridge et al. 2025). The weighted median method provides consistent estimates even when up to 50% of the genetic weight derives from invalid instruments (Bowden et al. 2016). The weighted mode method identifies the most frequently occurring causal effect estimate across clusters of variants, offering additional robustness to pleiotropy (Verbanck et al. 2018). MR Pleiotropy Residual Sum and Outlier (MR-PRESSO) detects and corrects for widespread horizontal pleiotropy by identifying and removing outlier variants before re-estimating the causal effect (Verbanck et al. 2018). In the present study, all five methods were applied within each ancestry group, and results were considered robust when effect estimates were directionally consistent across approaches and sensitivity analyses indicated no significant directional pleiotropy.

In the present study, our main objective was to investigate the potential causal relationship between COVID-19 and AMI across European, East Asian, South Asian, and African populations. To achieve this objective, we conducted a multi-ancestry two-sample Mendelian randomization (MR) analysis using ancestry-specific summary statistics from large-scale GWAS. COVID-19 exposure data were obtained from the COVID-19 Host Genetics Initiative Round 7 meta-analysis, while ancestry-matched AMI outcome data were obtained from the UK Biobank Whole-Genome Sequencing Consortium and Biobank Japan.

## 2 Methods

### 2.1 Study Design

We conducted a two-sample multi-ancestry MR analysis to evaluate the causal relationship between genetically predicted COVID-19 phenotypes and the risk of AMI (Figure 1) (Sekula et al. 2016; Sanderson et al. 2022; Burgess et al. 2019). MR uses genetic variants associated with an exposure as instrumental variables (IVs) to estimate causal effects while reducing susceptibility to residual confounding and reverse causation, provided that the instrumental-variable assumptions are satisfied (Sekula et al. 2016; Sanderson et al. 2022). We examined three COVID-19 phenotypes representing increasing disease severity as exposures: general COVID-19 susceptibility (infection), hospitalized COVID-19, and very severe respiratory confirmed COVID-19. To account for differences in genetic architecture and linkage disequilibrium (LD) patterns across populations, we performed analyses separately within four ancestry groups including European, East Asian, South Asian, and African.

**Figure 1.**
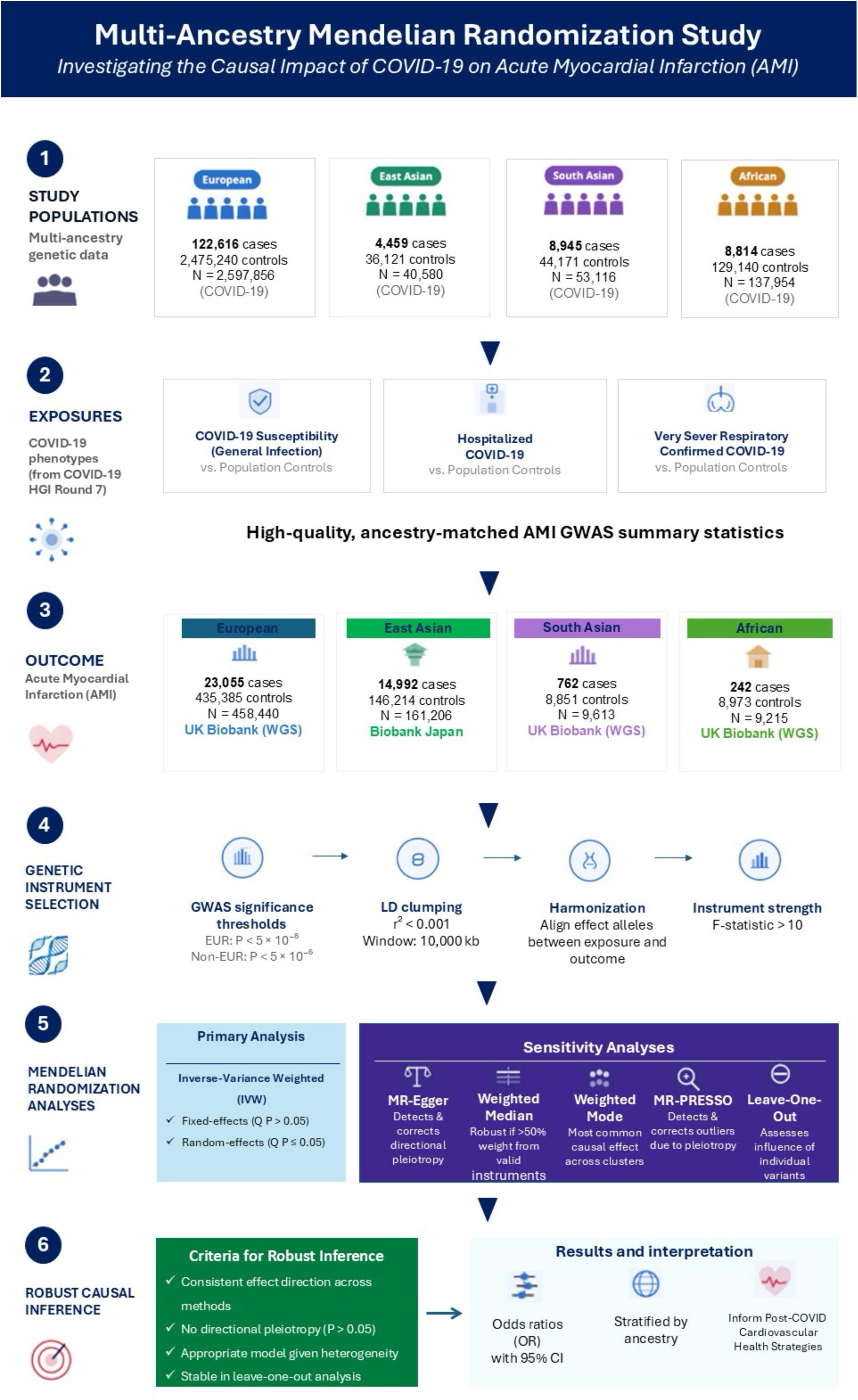
Study design and analytical framework of the multi-ancestry Mendelian randomization study. The study evaluated the potential causal effects of 3 COVID-19 phenotypes --- COVID-19 susceptibility (general infection), hospitalized COVID-19, and very severe respiratory confirmed COVID-19 --- on acute myocardial infarction (AMI) using ancestry-matched genome-wide association study summary statistics. Analyses were conducted separately in European, East Asian, South Asian, and African ancestry populations. Genetic instruments were selected using genome-wide significance thresholds of *P* < 5 × 10⁻⁸ for European ancestry and *P* < 5 × 10⁻⁶ for non-European ancestries, followed by linkage disequilibrium clumping, harmonization of effect alleles, and instrument-strength assessment. The primary Mendelian randomization analysis used inverse-variance weighting (IVW), with fixed-or random-effects models selected according to heterogeneity. Sensitivity analyses included MR-Egger, weighted median, weighted mode, MR-PRESSO, and leave-one-out analyses. Robust causal inference was based on consistency of effect direction across methods, absence of directional pleiotropy, appropriate modeling of heterogeneity, and stability in leave-one-out analyses. Effect estimates are reported as odds ratios (ORs) with 95% CIs and were stratified by ancestry.

### 2.2 Data Sources

We obtained GWAS summary statistics for the three COVID-19 phenotypes from the latest COVID-19 Host Genetics Initiative (HGI) Round 7 meta-analysis (released on April 8, 2022) (COVID-19 Host Genetics Initiative. 2022). This HGI provides large-scale GWAS summary statistics for general COVID-19 infection, hospitalized COVID-19, and very severe respiratory confirmed COVID-19 across multiple ancestral populations (Table 1). We similarly sourced outcome data for AMI from high-quality, population-level biobanks that match the ancestry groups of the exposure data. Specifically, AMI summary statistics for individuals of European, South Asian, and African ancestry were derived from the UK Biobank Whole-Genome Sequencing Consortium, while data for the East Asian population were obtained from Biobank Japan (UK Biobank 2025; Sakaue et al. 2021). These datasets rely on rigorous diagnostic criteria and large sample sizes to provide precise genetic estimates (Table 1). The integration of these diverse data sources into a multi-ancestry MR framework ensures that the identified causal relationships are both genetically reliable and globally representative.

**Table 1.**
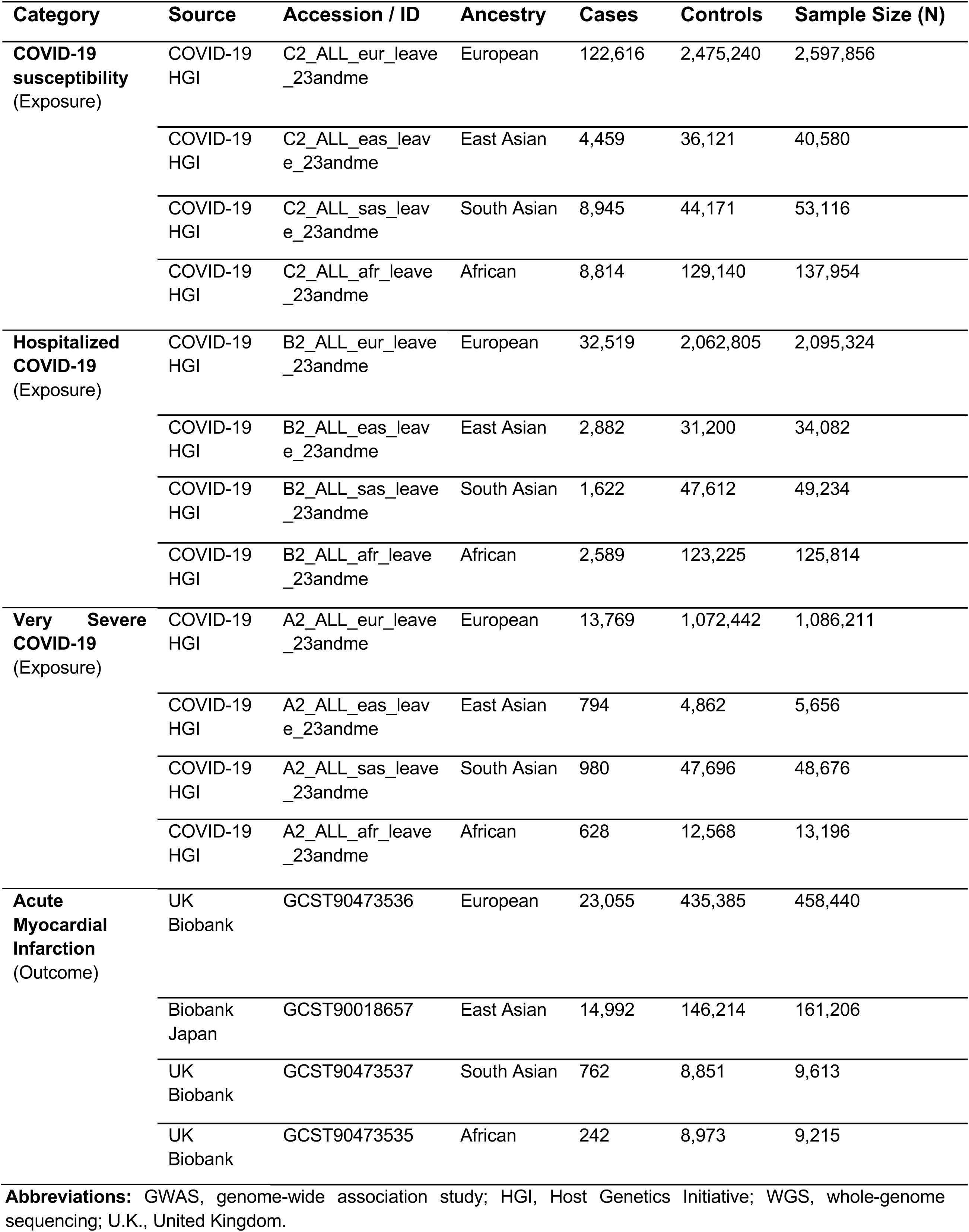
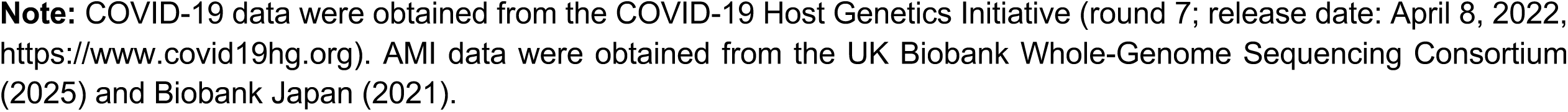
GWAS Summary Statistics Data Sources for COVID-19 and AMI.

| Category | Source | Accession / ID | Ancestry | Cases | Controls | Sample Size (N) |
| --- | --- | --- | --- | --- | --- | --- |
| **COVID-19 susceptibility**<br>(Exposure) | COVID-19 HGI | C2_ALL_eur_leave_23andme | European | 122,616 | 2,475,240 | 2,597,856 |
|  | COVID-19 HGI | C2_ALL_eas_leave_23andme | East Asian | 4,459 | 36,121 | 40,580 |
|  | COVID-19 HGI | C2_ALL_sas_leave_23andme | South Asian | 8,945 | 44,171 | 53,116 |
|  | COVID-19 HGI | C2_ALL_afr_leave_23andme | African | 8,814 | 129,140 | 137,954 |
| **Hospitalized COVID-19**<br>(Exposure) | COVID-19 HGI | B2_ALL_eur_leave_23andme | European | 32,519 | 2,062,805 | 2,095,324 |
|  | COVID-19 HGI | B2_ALL_eas_leave_23andme | East Asian | 2,882 | 31,200 | 34,082 |
|  | COVID-19 HGI | B2_ALL_sas_leave_23andme | South Asian | 1,622 | 47,612 | 49,234 |
|  | COVID-19 HGI | B2_ALL_afr_leave_23andme | African | 2,589 | 123,225 | 125,814 |
| **Very Severe COVID-19**<br>(Exposure) | COVID-19 HGI | A2_ALL_eur_leave_23andme | European | 13,769 | 1,072,442 | 1,086,211 |
|  | COVID-19 HGI | A2_ALL_eas_leave_23andme | East Asian | 794 | 4,862 | 5,656 |
|  | COVID-19 HGI | A2_ALL_sas_leave_23andme | South Asian | 980 | 47,696 | 48,676 |
|  | COVID-19 HGI | A2_ALL_afr_leave_23andme | African | 628 | 12,568 | 13,196 |
| **Acute Myocardial Infarction**<br>(Outcome) | UK Biobank | GCST90473536 | European | 23,055 | 435,385 | 458,440 |
|  | Biobank Japan | GCST90018657 | East Asian | 14,992 | 146,214 | 161,206 |
|  | UK Biobank | GCST90473537 | South Asian | 762 | 8,851 | 9,613 |
|  | UK Biobank | GCST90473535 | African | 242 | 8,973 | 9,215 |

**Abbreviations:** GWAS, genome-wide association study; HGI, Host Genetics Initiative; WGS, whole-genome sequencing; U.K., United Kingdom.
**Note:** COVID-19 data were obtained from the COVID-19 Host Genetics Initiative (round 7; release date: April 8, 2022, https://www.covid19hg.org). AMI data were obtained from the UK Biobank Whole-Genome Sequencing Consortium (2025) and Biobank Japan (2021).

### 2.3 Genetic instrument selection

We implemented a rigorous multistep screening protocol based on genome-wide association study (GWAS) summary statistics to select instrumental variables (IVs) for each COVID-19 phenotype. Genetic variants significantly associated with COVID-19 were selected using ancestry-specific genome-wide significance thresholds based on available sample sizes. Owing to the substantially larger sample size in European ancestry GWAS datasets, a stringent genome-wide significance threshold of *P* < 5 × 10⁻⁸ was applied. In contrast, for non-European ancestry populations with comparatively smaller sample sizes, a less stringent threshold of *P* < 5 × 10⁻⁶ was used to ensure adequate instrument strength and statistical power. To ensure independence among genetic instruments, Linkage disequilibrium (LD) clumping was performed using the *ieugwasr* R package within a threshold of r² < 0.001 within a 10,000-kb window. This clumping procedure was conducted separately within each ancestral population for every COVID-19 phenotype using the corresponding ancestry-specific reference panels.

We then performed data harmonization prior to analysis, to ensure correct allele alignment between exposure and outcome datasets. Proxy SNPs were not included in the analysis. To minimize weak-instrument bias, all retained genetic instruments demonstrated adequate strength, with F-statistics exceeding the conventional threshold of 10. Detailed characteristics of the selected IVs were provided in Supplementary Tables 1-12.

### 2.4 Mendelian Randomization (MR) Analysis

We implemented a comprehensive, multi-method MR framework to evaluate causal relationships between COVID-19 phenotypes and AMI risk. The inverse-variance weighted (IVW) method served as our primary analytical approach, conducted using both fixed-effects and multiplicative random-effects models, as it yields the most precise causal estimates when directional pleiotropy is absent (Bowden et al. 2016). Two IVW models were applied depending on the level of heterogeneity among genetic instruments: (1) Fixed-effects IVW, used when Cochran’s Q test indicated no heterogeneity (*P* > 0.05), (2) Multiplicative random-effects IVW, used when heterogeneity was detected (Bowden et al. 2018).

To enhance robustness and address potential violations of MR assumptions, we implemented three complementary estimation methods with distinct underlying assumptions. The MR-Egger method provides consistent causal effect estimates under the InSIDE (Instrument Strength Independent of Direct Effect) assumption and permits pleiotropy bias estimation through the intercept (Burgess et al. 2017). The weighted median method offers robustness to invalid instruments, providing consistent estimates when at least 50% of the weight in the analysis derives from valid instruments (Bowden et al. 2016). The weighted mode method identifies the most frequently occurring causal effect estimate among clusters of genetic variants, thereby providing protection against pleiotropy (Burgess et al. 2020).

All analyses were stratified by ancestry (European, East Asian, South Asian, and African) to account for population-specific genetic architecture and linkage disequilibrium patterns. Causal effects of binary COVID-19 exposures on AMI risk were reported as odds ratios (ORs) with corresponding 95% confidence intervals.

### 2.5 Heterogeneity and Sensitivity Analyses

The validity of MR analyses relies on three core assumptions: (1) Relevance: genetic variants must show a strong and statistically reliable association with the exposure, typically evidenced by an F-statistic greater than 10, which indicates adequate instrument strength, (2) Independence: genetic variants are independent of confounder, and (3) Exclusion restriction: genetic variants affect the outcome exclusively through the exposure. Violation of these assumptions, particularly through horizontal pleiotropy whereby instrumental variables affect the outcome through pathways independent of the exposure, can introduce bias into causal estimates. To evaluate potential violations of these assumptions, several sensitivity analyses were conducted.

### Heterogeneity Assessment

Between-variant heterogeneity was quantified using Cochran’s Q statistic for both IVW and MR-Egger models. Substantial heterogeneity may indicate pleiotropy, population stratification, or other violations of instrument validity. As noted above, heterogeneity test results determined model selection, with *P* > 0.05 supporting fixed-effects models and *P* ≤ 0.05 necessitating random-effects approaches to account for excess variance.

### Horizontal Pleiotropy Assessment

Multiple approaches were employed to detect and account for pleiotropic effects. The MR-Egger intercept test formally evaluates directional pleiotropy, with significant deviation from zero (*P* < 0.05) indicating its presence. Additionally, we applied the MR-PRESSO global test to identify outlier variants exhibiting pleiotropic effects and to obtain bias-corrected estimates following the removal of outlier pleiotropic variants (Verbanck et al. 2018).

### Individual Variant Influence

To evaluate whether any single variant disproportionately influences overall results, we performed leave-one-out sensitivity analysis by sequentially removing each SNP and recalculating the IVW estimate to assess the stability of causal inferences.

### 2.6 Criteria for Robust Causal Inference

Results were considered robust and indicative of genuine causal relationships when the following criteria were satisfied: (1) consistency of effect estimates across multiple analytical methods (IVW, weighted median, MR-Egger, weighted mode) (2) absence of directional pleiotropy (absence of directional pleiotropy, as indicated by a non-significant MR-Egger intercept (*P* > 0.05) (3) acceptable levels of heterogeneity with appropriate model selection, and (4) stability in leave-one-out sensitivity analysis. Convergent evidence across these complementary validation approaches strengthens confidence in causal relationships between COVID-19 infection and AMI risk.

### 2.7 Statistical analysis

All MR analyses were implemented using the TwoSampleMR and MR-PRESSO packages in R software (version 4.1.2). A two-tailed *P*-value < 0.05 was considered statistically significant unless otherwise specified. Given that the five MR methods applied represent complementary approaches to estimating the same causal parameter rather than independent hypothesis tests, no false discovery rate correction was applied across methods. Causal inference was based on consistency of effect direction and magnitude across approaches.

## 3 Results

We conducted a comprehensive MR analysis to investigate the causal relationship between COVID-19 severity and AMI risk across multiple ancestral populations. Three COVID-19 severity phenotypes were examined: COVID-19 infection versus population, hospitalized COVID-19, and very severe respiratory confirmed COVID-19 versus population. The analysis was performed separately for European, East Asian, South Asian, and African ancestral groups.

### 3.1 European Ancestry Population

In European ancestry, MR analyses did not identify statistically significant causal associations between genetically predicted COVID-19 susceptibility and AMI across any MR methods. The IVW-fixed effects model yielded an odds ratio (OR) of 0.92 (95% CI: 0.82-1.04, *P* = 0.182). Cochran’s Q test showed no evidence of excess heterogeneity (*P* = 0.840). Complementary MR methods, including the weighted median, weighted mode, and MR-Egger approaches, yielded consistent null findings (Figure 2). Sensitivity analyses revealed no evidence of heterogeneity or directional pleiotropy for genetically predicted COVID-19 susceptibility (Table 2).

**Figure 2.**
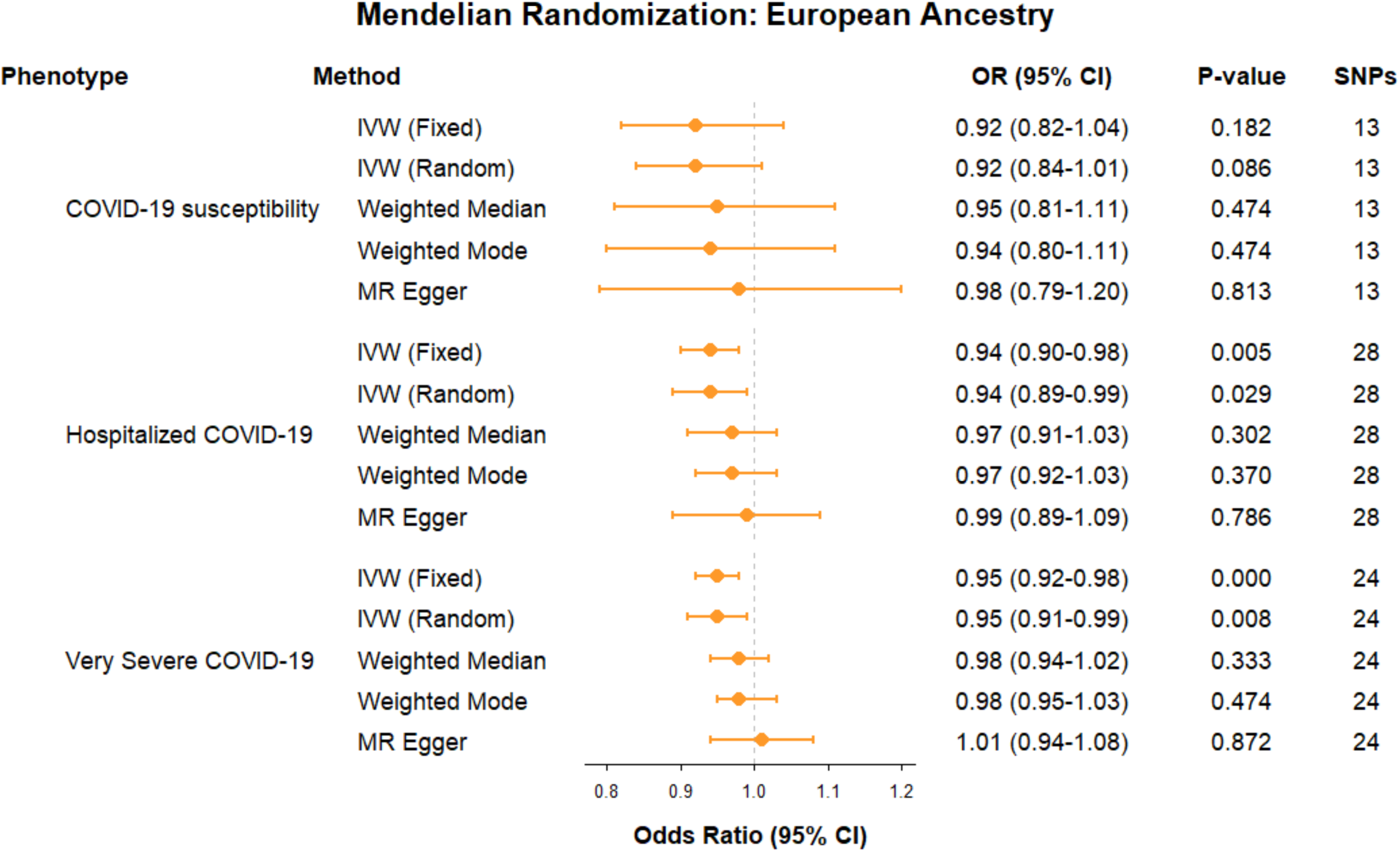
Mendelian randomization estimates for the association of COVID-19 phenotypes with acute myocardial infarction in European ancestry. Forest plots show odds ratios (ORs) and 95% CIs for the associations of genetically predicted COVID-19 infection, hospitalized COVID-19, and very severe COVID-19 with acute myocardial infarction using inverse-variance weighting (IVW), weighted median, weighted mode, and MR-Egger methods. The vertical dashed line represents the null association (OR=1.00). Each point represents the OR, and horizontal lines represent the corresponding 95% CI. The number of single-nucleotide polymorphisms (SNPs) included in each analysis and 2-sided P values are shown. For IVW, fixed-and random-effects estimates are presented according to the prespecified heterogeneity criteria.

**Table 2.** Horizontal Pleiotropy and Heterogeneity analyses of MR causal associations between COVID-19 phenotypes and AMI across ancestral groups.

| Ancestry | COVID-19 Exposure Type | MR-Egger Q<br>(*P*-value) | IVW Q<br>(*P*-value) | MR-Egger Intercept<br>(*P*-value) |
| --- | --- | --- | --- | --- |
| European | COVID-19 vs. population | 6.82 (0.813) | 7.26 (0.840) | −0.0036 (0.521) |
|  | Hospitalized COVID-19 vs. population | 41.78 (0.026) | 43.65 (0.022) | −0.0057 (0.291) |
|  | Very severe COVID-19 vs. population | 33.91 (0.050) | 40.15 (0.015) | −0.0113 (0.057) |
| East Asian | COVID-19 vs. population | 5.27 (0.383) | 6.85 (0.335) | 0.0361 (0.275) |
|  | Hospitalized COVID-19 vs. population | 6.77 (0.453) | 10.12 (0.257) | −0.0308 (0.110) |
|  | Very severe COVID-19 vs. population | 4.15 (0.763) | 4.17 (0.841) | 0.0026 (0.875) |
| South Asian | COVID-19 vs. population | 6.92 (0.906) | 6.92 (0.938) | −0.0025 (0.959) |
|  | Hospitalized COVID-19 vs. population | 7.71 (0.957) | 7.72 (0.972) | −0.0042 (0.935) |
|  | Very severe COVID-19 vs. population | 6.19 (0.992) | 7.09 (0.989) | −0.0422 (0.356) |
| African | COVID-19 vs. population | 31.78 (0.164) | 37.69 (0.065) | 0.1498 (0.041) |
|  | Hospitalized COVID-19 vs. population | 21.37 (0.672) | 25.14 (0.511) | −0.1453 (0.064) |
|  | Very severe COVID-19 vs. population | 10.56 (0.228) | 12.20 (0.202) | −0.1397 (0.297) |

**Notes:** Heterogeneity was assessed using Cochran’s Q statistic for MR-Egger and IVW methods. Horizontal pleiotropy was evaluated using the MR-Egger intercept test. Significant heterogeneity or pleiotropy was defined as $P < 0.05$ . All analyses were adjusted for ancestry-specific genetic instruments and summary-level data from publicly available GWAS summary statistics. **Abbreviations:** IVW, inverse variance weighted; MR-PRESSO, Mendelian Randomization Pleiotropy Residual Sum and Outlier.

As significant heterogeneity was detected by Cochran’s Q for hospitalized COVID-19 (MR-Egger Q = 41.78, P = 0.026; IVW Q = 43.65, *P* = 0.022) and very severe respiratory confirmed COVID-19 (MR-Egger Q = 33.91, *P* = 0.050; IVW Q = 40.15, *P* = 0.015), the IVW random-effects model was used as the primary estimator for these two phenotypes to account for between-instrument variability. Genetically predicted hospitalized COVID-19 showed a nominally significant inverse association with AMI in the IVW random-effects analysis (OR = 0.94, 95% CI: 0.89-0.99, *P* = 0.029). Sensitivity analyses using weighted median, weighted mode, and MR-Egger methods produced directionally consistent but non-significant estimates (Figure 2).

Additionally, genetically predicted very severe respiratory confirmed COVID-19 showed a statistically significant negative association with AMI risk in the IVW-random effects model (OR = 0.95, 95% CI: 0.91-0.99, *P* = 0.008). Directionally consistent but non-significant results were observed using the weighted median, weighted mode, and MR-Egger methods, indicating consistent effect direction but limited statistical support across all MR approaches (Figure 2).

MR-Egger intercept tests did not indicate significant directional pleiotropy for either hospitalized or very severe COVID-19 (both *P* > 0.05, Table 2). MR-PRESSO tests indicated no outlier variants were identified in both groups. We further demonstrated that the overall estimates were not driven by individual SNPs and showed no substantial horizontal pleiotropy, supporting the robustness of these findings (Supplementary Figures 1-3).

### 3.2 East Asian Ancestry Population

In the East Asian population, we found no statistically significant evidence supporting a causal relationship between genetically predicted COVID-19 susceptibility and AMI risk. For general COVID-19 susceptibility, the IVW fixed-effects estimate yielded an OR of 0.97 (95% CI: 0.91-1.03; *P* = 0.349), with consistent null findings across weighted-median (OR = 0.98, 95% CI: 0.89-1.07; *P* = 0.604), weighted-mode (OR = 1.02, 95% CI: 0.88-1.19; *P* = 0.787), and MR-Egger (OR = 0.86, 95% CI: 0.70-1.06; *P* = 0.205) methods. Similarly, for hospitalized COVID-19, the IVW fixed-effects estimate was 0.99 (95% CI: 0.94-1.04; *P* = 0.711), and for very severe COVID-19 the corresponding estimate was 0.98 (95% CI: 0.95-1.01; *P* = 0.182). All complementary MR methods yielded concordant non-significant results for both severe phenotypes, as illustrated in Figure 3.

**Figure 3.**
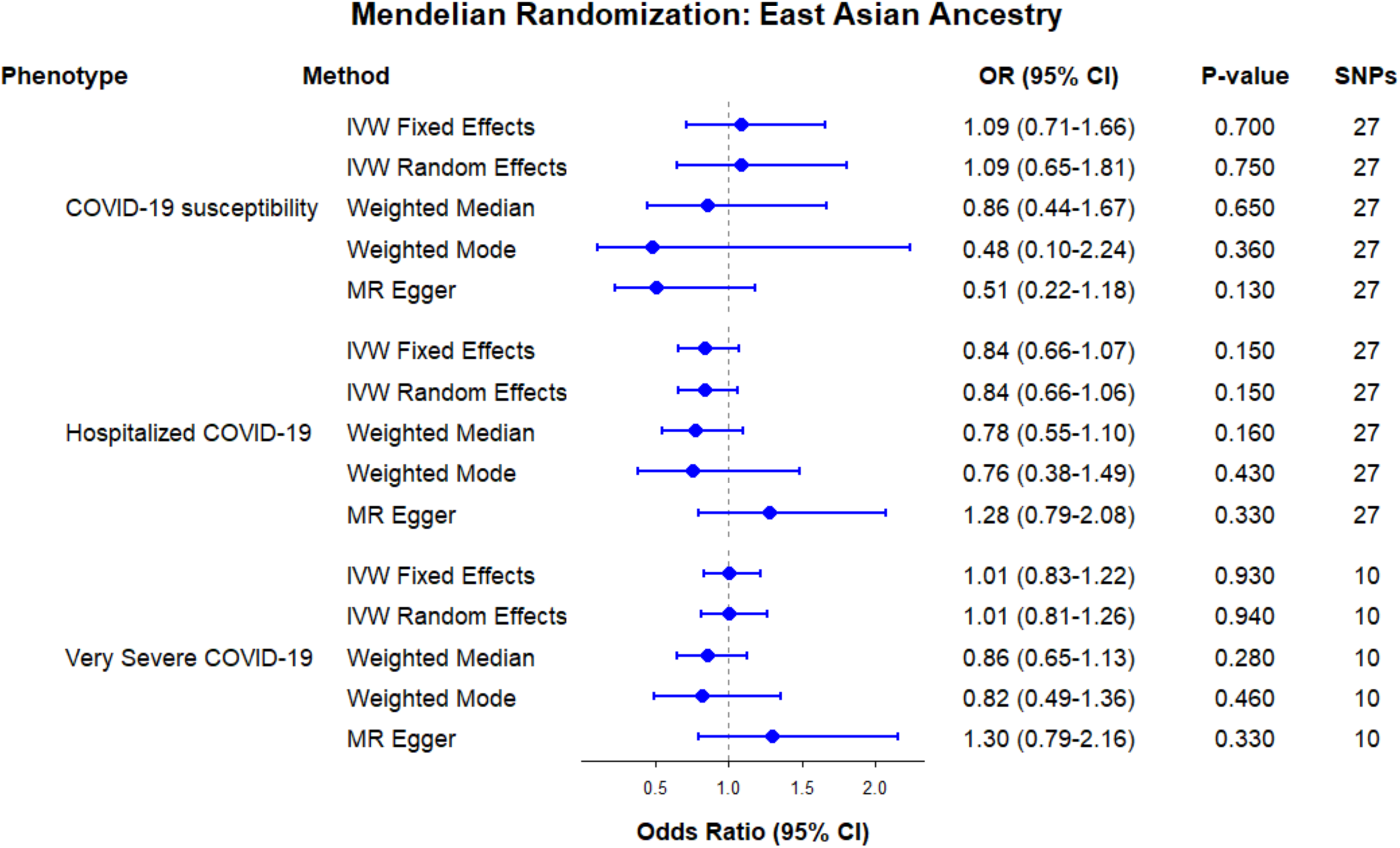
Mendelian randomization estimates for the association of COVID-19 phenotypes with acute myocardial infarction in East Asian ancestry. Forest plots show odds ratios (ORs) and 95% CIs for the associations of genetically predicted COVID-19 infection, hospitalized COVID-19, and very severe COVID-19 with acute myocardial infarction using inverse-variance weighting (IVW), weighted median, weighted mode, and MR-Egger methods. The vertical dashed line represents the null association (OR=1.00). Each point represents the OR, and horizontal lines represent the corresponding 95% CI. The number of single-nucleotide polymorphisms (SNPs) included in each analysis and 2-sided P values are shown. For IVW, fixed-and random-effects estimates are presented according to the prespecified heterogeneity criteria.

Prior to analysis of hospitalized COVID-19 (B2), MR-PRESSO identified 1 outlier SNP (rs11066150), which was removed, leaving 9 instruments for that phenotype. Sensitivity analyses showed no evidence of heterogeneity or horizontal pleiotropy (Table 2), supported by leave-one-out, scatter, and funnel plots (Supplementary Figures 4-6). This data indicates that genetic liability to COVID-19, across a spectrum of infection severity, does not appear to confer an elevated risk of AMI in East Asian ancestries. These findings are consistent with other Mendelian randomization studies reporting no clear causal association between genetically predicted COVID-19 susceptibility and cardiovascular outcomes (Wang et al. 2022).

### 3.3 South Asian Ancestry Population

We found that MR analysis in South Asian populations revealed no consistent evidence of a causal association between genetically predicted COVID-19 susceptibility and AMI risk. Cochran’s Q test showed no evidence of excess heterogeneity (Table 2). For general COVID-19 susceptibility, the primary IVW fixed-effects estimate yielded an OR of 0.93 (95% CI: 0.68-1.27; *P* = 0.636), with consistent null findings across all complementary methods (weighted-median OR = 0.93, *P* = 0.712; weighted-mode OR = 0.94, *P* = 0.900; MR-Egger OR = 0.94, *P* = 0.883). For hospitalized COVID-19, the fixed-effects IVW estimate was used for primary interpretation. No statistically significant association with AMI was observed (OR, 0.92; 95% CI, 0.82–1.02; *P* = 0.126). The weighted median, weighted mode, and MR-Egger estimates were also nonsignificant (Figure 4). For very severe COVID-19, the IVW fixed-effects estimate was 1.02 (95% CI: 0.94-1.12; *P* = 0.587), again with no significant associations across sensitivity analyses (as shown in Figure 4).

**Figure 4.**
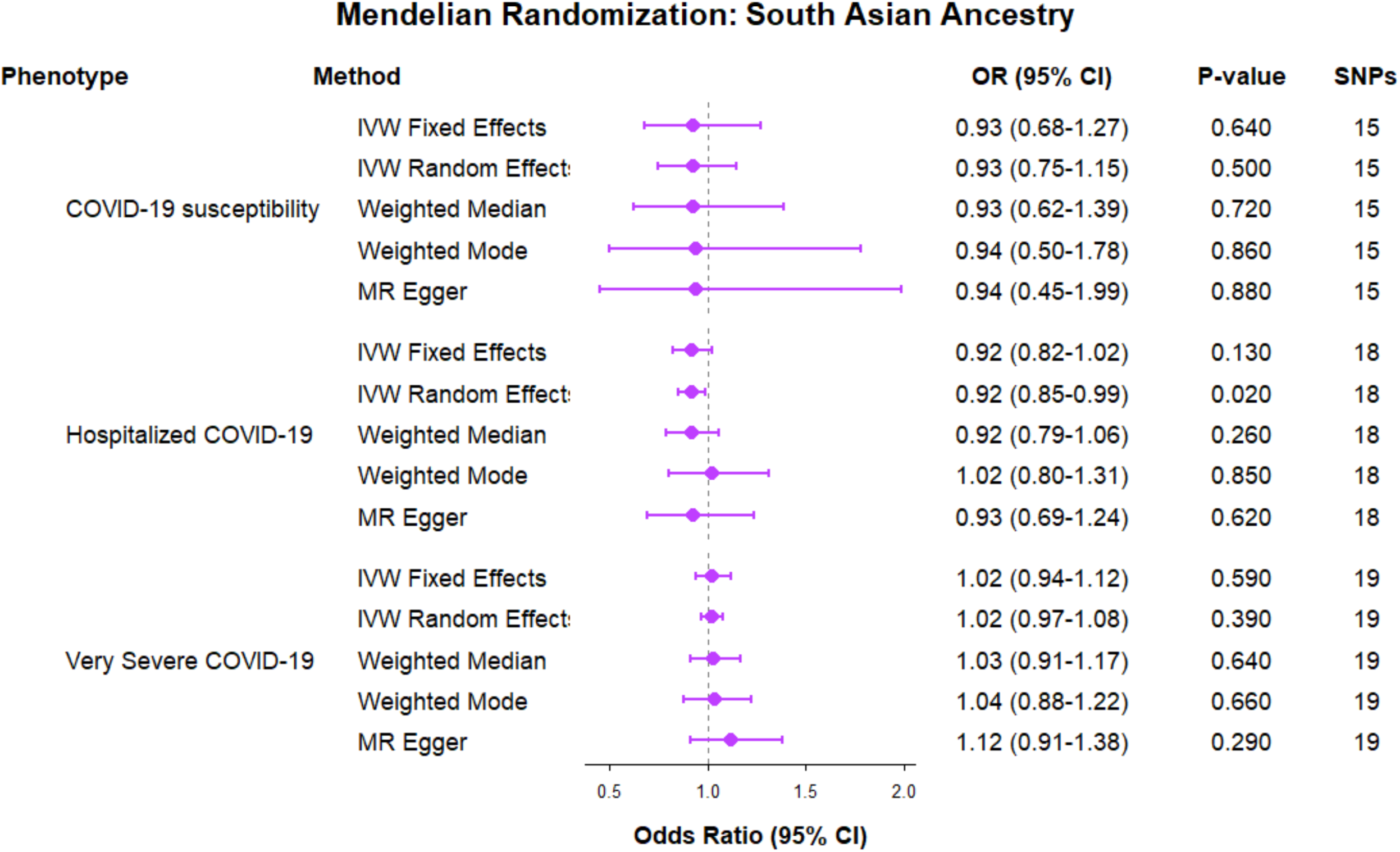
Mendelian randomization estimates for the association of COVID-19 phenotypes with acute myocardial infarction in South Asian ancestry. Forest plots show odds ratios (ORs) and 95% CIs for the associations of genetically predicted COVID-19 infection, hospitalized COVID-19, and very severe COVID-19 with acute myocardial infarction using inverse-variance weighting (IVW), weighted median, weighted mode, and MR-Egger methods. The vertical dashed line represents the null association (OR=1.00). Each point represents the OR, and horizontal lines represent the corresponding 95% CI. The number of single-nucleotide polymorphisms (SNPs) included in each analysis and 2-sided P values are shown. For IVW, fixed-and random-effects estimates are presented according to the prespecified heterogeneity criteria.

Heterogeneity and pleiotropy tests were non-significant (Table 2), with sensitivity plots confirming consistency (Supplementary Figures 7-9). Collectively, these findings suggest that genetic predisposition to COVID-19 does not translate into a materially increased risk of AMI in South Asian populations, aligning with the null results observed in East Asian ancestries.

### 3.4 African Ancestry Population

In African ancestry populations, we found that MR analyses indicated no statistically significant causal relationship between genetically predicted COVID-19 susceptibility and AMI risk. For general COVID-19 susceptibility, the IVW fixed-effects estimate yielded an OR of 1.09 (95% CI: 0.71-1.66; *P* = 0.696), with consistent non-significant results across weighted-median (OR = 0.86, 95% CI: 0.45-1.62; *P* = 0.634), weighted-mode (OR = 0.48, 95% CI: 0.09-2.55; *P* = 0.394), and MR-Egger (OR = 0.51, 95% CI: 0.22-1.18; *P* = 0.128) methods.

For hospitalized COVID-19, the OR by IVW fixed-effects estimate was 0.84 (95% CI: 0.66-1.07; *P* = 0.155), with no significant associations across sensitivity methods (weighted-median OR = 0.78, *P* = 0.163; weighted-mode OR = 0.76, *P* = 0.441; MR-Egger OR = 1.28, *P* = 0.334). For very severe COVID-19, the OR of IVW fixed-effects estimate was 1.01 (95% CI: 0.83-1.22; *P* = 0.928), and all sensitivity analyses similarly showed no significant associations (weighted-median OR = 0.86, *P* = 0.270; weighted-mode OR = 0.82, *P* = 0.667; MR-Egger OR = 1.30, *P* = 0.331) (Figure 5).

**Figure 5.**
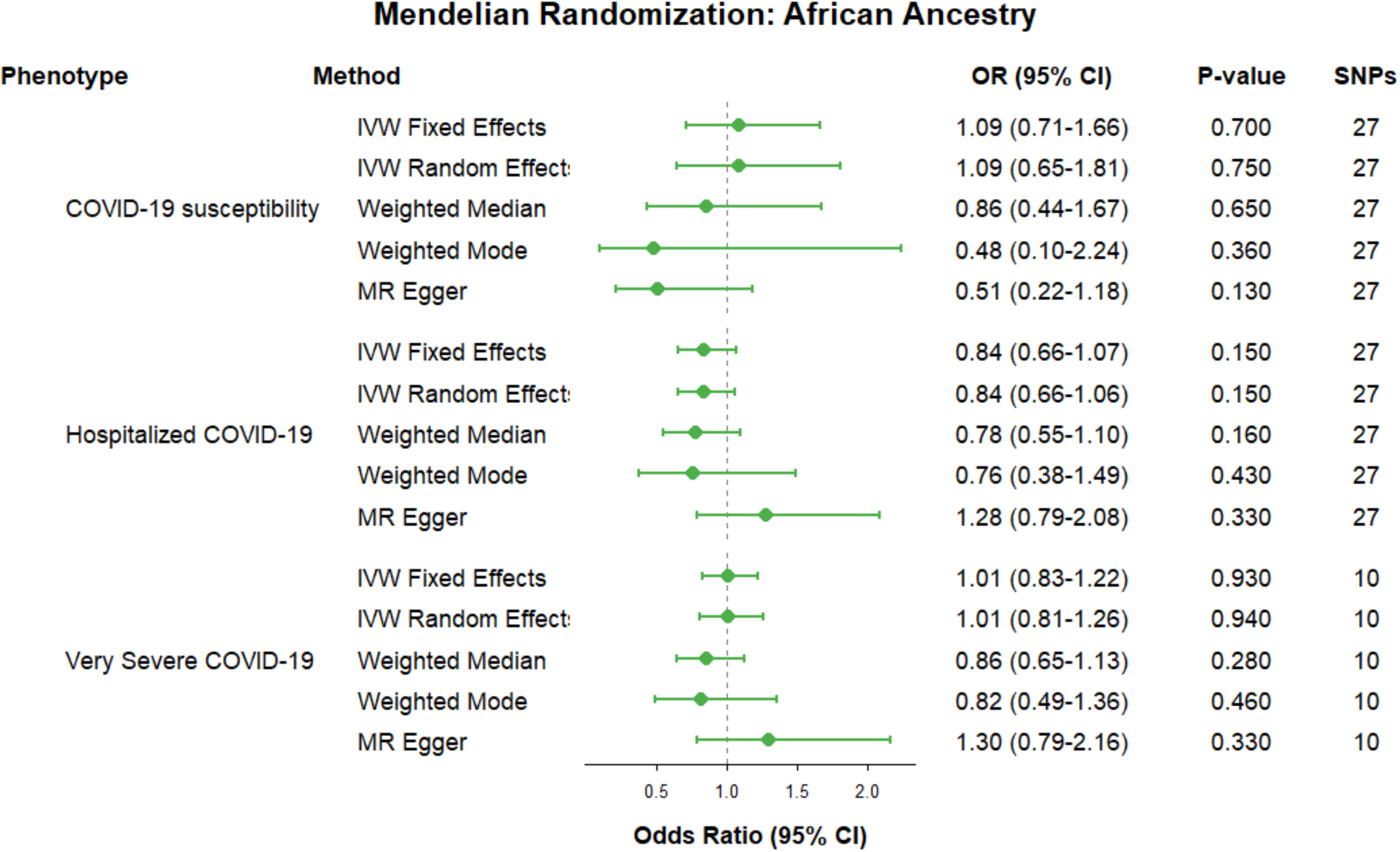
Mendelian randomization estimates for the association of COVID-19 phenotypes with acute myocardial infarction in African ancestry. Forest plots show odds ratios (ORs) and 95% CIs for the associations of genetically predicted COVID-19 infection, hospitalized COVID-19, and very severe COVID-19 with acute myocardial infarction using inverse-variance weighting (IVW), weighted median, weighted mode, and MR-Egger methods. The vertical dashed line represents the null association (OR=1.00). Each point represents the OR, and horizontal lines represent the corresponding 95% CI. The number of single-nucleotide polymorphisms (SNPs) included in each analysis and 2-sided P values are shown. For IVW, fixed-and random-effects estimates are presented according to the prespecified heterogeneity criteria.

Notably, the MR-Egger intercept test indicated significant directional pleiotropy for general COVID-19 susceptibility in African ancestry (*P* = 0.041), suggesting that the IVW estimate for this phenotype should be interpreted with caution. Other sensitivity test plots were shown in Supplementary Figures 10-12.

## 4 Discussion

We conducted a comprehensive multi-ancestry Mendelian randomization (MR) study using COVID-19 Host Genetics Initiative Round 7 (April 2022) GWAS summary statistics as exposure data and ancestry-specific AMI GWAS from UK Biobank and Biobank Japan as outcome data, to investigate the causal relationship between genetic liability to COVID-19 susceptibility or severity and acute AMI risk across European, East Asian, South Asian, and African populations. We identified nominal inverse associations between genetic liability to hospitalized and very severe COVID-19 and AMI risk in European ancestry, and a suggestive but non-significant inverse association for hospitalized COVID-19 in South Asian ancestry under the pre-specified fixed-effects model. However, these associations were not consistently supported by complementary MR methods. No statistically significant associations were identified in the East Asian or African ancestry populations.

These findings support a complex relationship between COVID-19 and AMI and are broadly consistent with recent two-sample Mendelian randomization studies, which have not identified a strong causal link between COVID-19 phenotypes and AMI or related cardiovascular outcomes, despite persistent observational evidence of elevated risk (Wang et al. 2022; Zheng et al. 2023). The lack of consistent causal signals across ancestries, phenotypes, and MR methods suggests that genetic liability to COVID-19 susceptibility or severity alone may not explain the observed clinical association between COVID-19 and AMI. Ethnic disparities in COVID-19 severity and outcomes further emphasize the importance of non-genetic factors on myocardial infarction risk (Magesh et al. 2021; Coughlin and Young 2020). Observational studies have demonstrated substantially higher mortality and complication rates among Asian, Black, and Hispanic populations compared with European-ancestry individuals, despite heterogeneous infection and mortality patterns worldwide (Prabhakaran et al. 2022). Motivated by these ethnic disparities, our multi-ancestry MR analysis shows that ancestry-specific genetic liability to COVID-19 is unlikely to drive excess AMI risk across populations. Instead, social determinants of health, comorbidity burden, and access to care likely predominate (Sundermeyer et al. 2025; Erskine et al. 2018; Del Prete et al. 2022).

Multiple biological mechanisms plausibly link COVID-19 to myocardial injury and infarction. These include direct effects, such as viral invasion of cardiomyocytes and endothelial cells via angiotensin-converting enzyme 2 (ACE2) receptors, resulting in localized cellular damage, as well as indirect pathways, including cytokine storm-mediated systemic inflammation, prothrombotic states leading to coronary microthrombosis, myocardial oxygen supply-demand mismatch secondary to hypoxia and respiratory failure, and coronary vasospasm (Canale et al. 2022; Zanza et al. 2022; Abou-Ismail et al. 2020; McFadyen et al. 2020; Esposito et al. 2021; Maitz et al. 2023; Chidambaram et al. 2024). These processes can contribute to both type 1 and type 2 myocardial infarction, whereby myocardial damage may result from direct viral invasion or indirect mechanisms including systemic inflammation, immune-mediated response, and renin-angiotensin system dysregulation, or from an imbalance between myocardial oxygen supply and demand and are associated with increased mortality and adverse cardiovascular outcomes (Gupta et al. 2017). Emerging experimental evidence has also suggested that the SARS-CoV-2 spike protein may affect vascular endothelial function independently of active viral replication, although the clinical relevance of these findings requires further validation in human studies (Gultom et al. 2025; Imig 2022).

Overall, our results show no consistent evidence that genetic susceptibility to or severity of COVID-19 causally increases the risk of AMI across different ancestries. Notably, this study addresses a major literature gap by evaluating population-specific genetic data for East Asian, South Asian, and African ancestries. Despite heterogeneity in European cohorts, results remained consistent across complementary MR sensitivity models. While modest inverse associations were observed for genetically predicted severe and hospitalized COVID-19 in European ancestry, and a suggestive but non-significant inverse association was observed for hospitalized COVID-19 in South Asian ancestry under the pre-specified analytical framework, these findings were not uniform across ancestries or MR sensitivity methods, suggesting limited generalizability.

Several limitations warrant consideration. First, there were uneven sample sizes that might reduce statistical power in smaller non-European cohorts, though consistent null results in modest size of East Asian populations support overall robustness. Additionally, because MR analysis reflects lifelong genetic liability rather than acute infection dynamics, our null genetic findings contrast with observational data showing short-term cardiovascular risk during active illness. Finally, unmeasured extrinsic factors like vaccination, treatments, and comorbidities emphasize the need to investigate non-genetic mechanisms driving post-COVID cardiovascular outcomes.

## Conclusion

While we found significant inverse and potentially causal associations between genetic liability to hospitalized and very severe COVID-19 and AMI risk in European ancestry, neither finding was robust across all sensitivity analyses, suggesting the causal association was not robust. In South Asian ancestry, hospitalized COVID-19 showed a suggestive but non-significant inverse association under the pre-specified fixed-effects model. No significant causal associations were identified for any COVID-19 phenotype in East Asian or African ancestries. These findings highlight the complexity of the COVID-19 and AMI association and shed light on potential strategies to promote post-COVID cardiovascular health.

## Data Availability

The study used (or will use) ONLY openly available human data that were originally located at COVID-19 Host Genetics Initiative (COVID-19 HGI), Release 7 (https://www.covid19hg.org/results/r7/), including phenotypes A2 (very severe respiratory confirmed COVID-19), B2 (hospitalized COVID-19), and C2 (COVID-19 infection). Acute myocardial infarction (AMI) GWAS summary statistics were obtained from the NHGRI-EBI GWAS Catalog: European ancestry: https://www.ebi.ac.uk/gwas/studies/GCST90473536,
East Asian ancestry: https://www.ebi.ac.uk/gwas/studies/GCST90018657,
South Asian ancestry: https://www.ebi.ac.uk/gwas/studies/GCST90473537,
African ancestry: https://www.ebi.ac.uk/gwas/studies/GCST90473535. The 1000 Genomes Phase 3 reference panel used for linkage disequilibrium clumping was publicly available at http://fileserve.mrcieu.ac.uk/ld/1kg.v3.tgz.

## Acknowledgements

The authors thank all participants and investigators from the COVID-19 Host Genetics Initiative (Round 7) for making the GWAS summary statistics publicly available, without which this multi-ancestry analysis would not have been possible. We also gratefully acknowledge the participants and researchers of the UK Biobank Whole-Genome Sequencing Consortium and Biobank Japan for providing the ancestry-matched acute myocardial infarction summary statistics.

## Ethics Declarations

This study utilized publicly available genome-wide association study (GWAS) summary statistics from the COVID-19 Host Genetics Initiative (Round 7), the UK Biobank Whole-Genome Sequencing Consortium, and Biobank Japan. No new human participants were recruited, no individual-level data were accessed or analyzed, and no primary data collection occurred.

## Availability of data and materials

The datasets analyzed in the current study are publicly available summary statistics. COVID-19 genetic association data were obtained from the COVID-19 Host Genetics Initiative (COVID-19-HGI) release 7 (https://www.covid19hg.org/results/r7/), including phenotypes A2 (very severe respiratory confirmed COVID-19), B2 (hospitalized COVID-19), and C2 (COVID-19 infection), across European, East Asian, South Asian, and African ancestries. AMI GWAS summary statistics were retrieved from the NHGRI-EBI GWAS Catalog (https://www.ebi.ac.uk/gwas/):

- European: https://www.ebi.ac.uk/gwas/studies/GCST90473536
- East Asian: https://www.ebi.ac.uk/gwas/studies/GCST90018657
- South Asian: https://www.ebi.ac.uk/gwas/studies/GCST90473537
- African: https://www.ebi.ac.uk/gwas/studies/GCST90473535

LD clumping was performed using the 1000 Genomes Phase 3 reference panel, available at: http://fileserve.mrcieu.ac.uk/ld/1kg.v3.tgz.

The analytic code can be found at https://github.com/Katherine-Zhou-git/MultiAncestry-MR-AMI.

## Author Contributions

Study concept and research elaboration: Katherine Zhou and Ying Ma. Data statistical analysis and draft the initial manuscript: Katherine Zhou, Interpreted the results and contributed to manuscript writing, revision, and final approval: Katherine Zhou and Ying Ma. Study supervision: Ying Ma.

## Non-standard Abbreviations and Acronyms

AFR: African (ancestry)
AMI: Acute Myocardial Infarction
BBJ: Biobank Japan (for East Asian AMI data)
CI: Confidence Interval
COVID-19: Coronavirus Disease 2019
EUR: European (ancestry)
EAS: East Asian (ancestry)
GWAS: Genome-Wide Association Study
HGI: COVID-19 Host Genetics Initiative
IVW: Inverse-Variance Weighted
LD: Linkage Disequilibrium
MR: Mendelian Randomization
MR-Egger: Mendelian Randomization Egger method
MR-PRESSO: Mendelian Randomization Pleiotropy RESidual Sum and Outlier
OR: Odds Ratio
P: P-value
Q: Cochran’s Q statistic (for heterogeneity)
SAS: South Asian (ancestry)
SNP: Single Nucleotide Polymorphism
UK Biobank: United Kingdom Biobank (for European, South Asian, African AMI data)
WGS: Whole-Genome Sequencing (UK Biobank consortium)

