## Supplementary Information for "Investigating the Causal Impact of COVID-19 on Acute Myocardial Infarction: A Multi-ancestry Mendelian Randomization Study"

### Supplementary Figures

**Supplementary Figure 1.** MR analyses evaluating the causal association between COVID-19 infection and AMI in European ancestry.

A

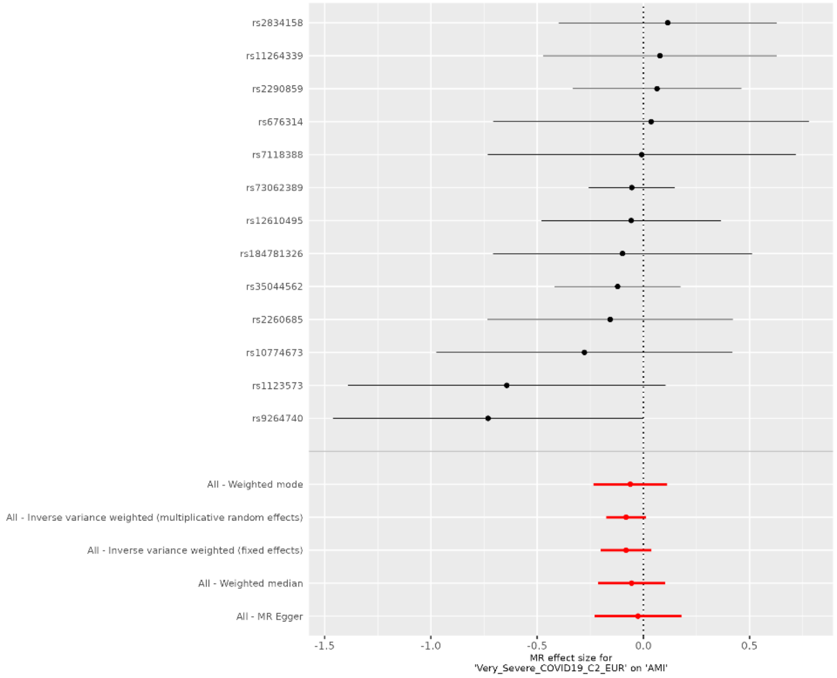


B

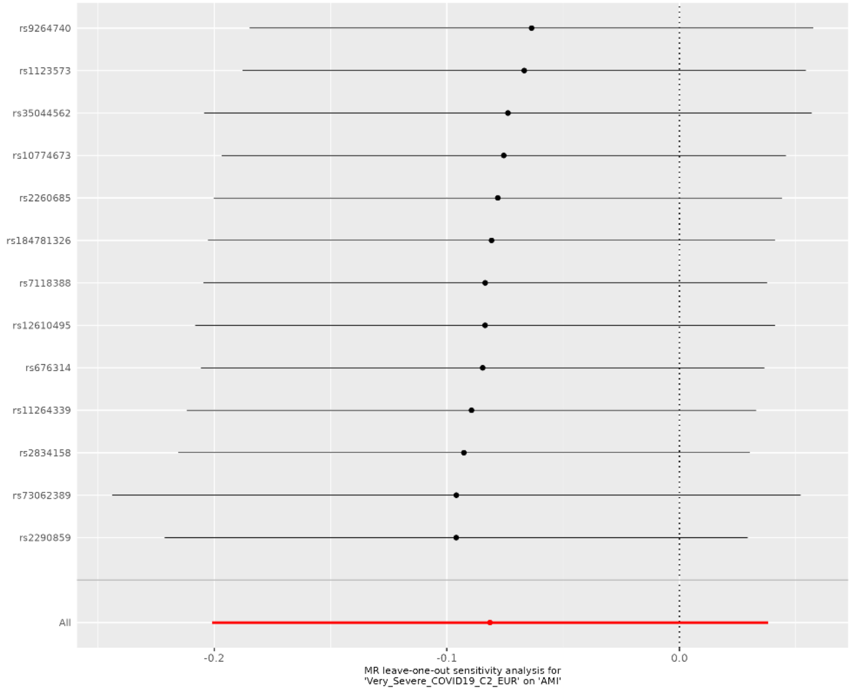

C

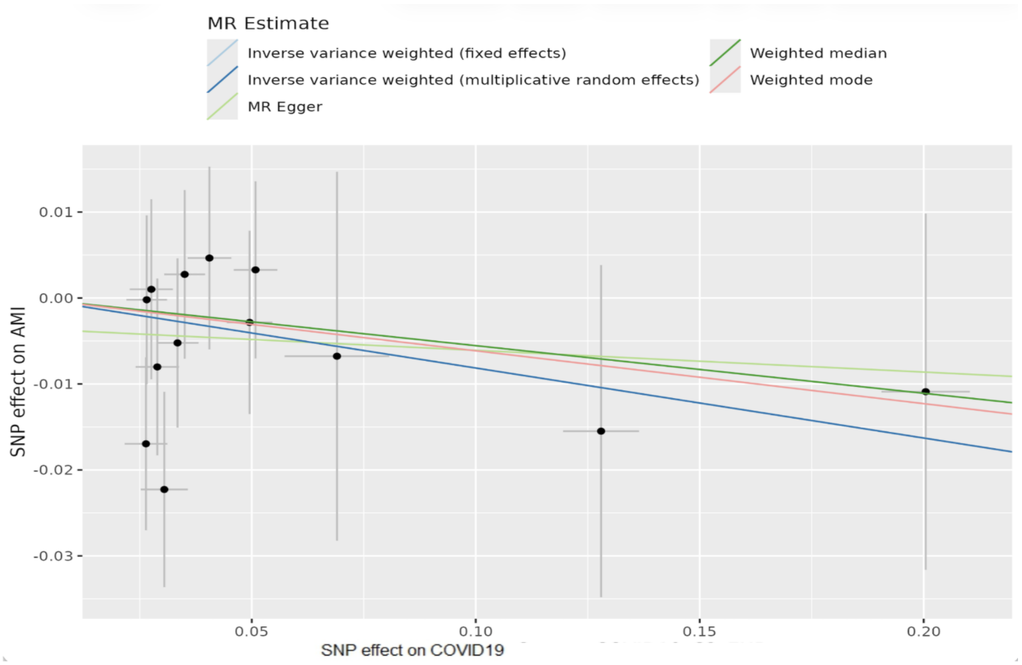


D

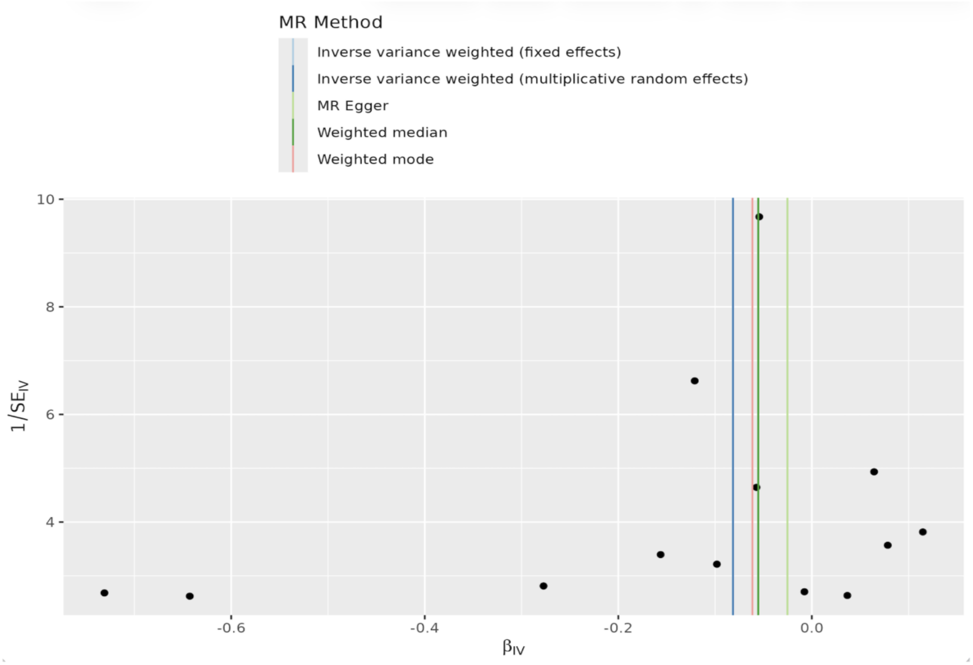


(A) Forest plot of individual SNP causal estimates. (B) Leave-one-out sensitivity analysis. (C) Scatter plot of SNP effects on COVID-19 and AMI with MR regression lines. (D) Funnel plot assessing instrument heterogeneity and potential pleiotropy. Abbreviations: AMI, acute myocardial infarction; MR, Mendelian randomization.

**Supplementary Figure 2.** MR analyses evaluating the causal association between hospitalized COVID-19 and AMI in European ancestry.

A

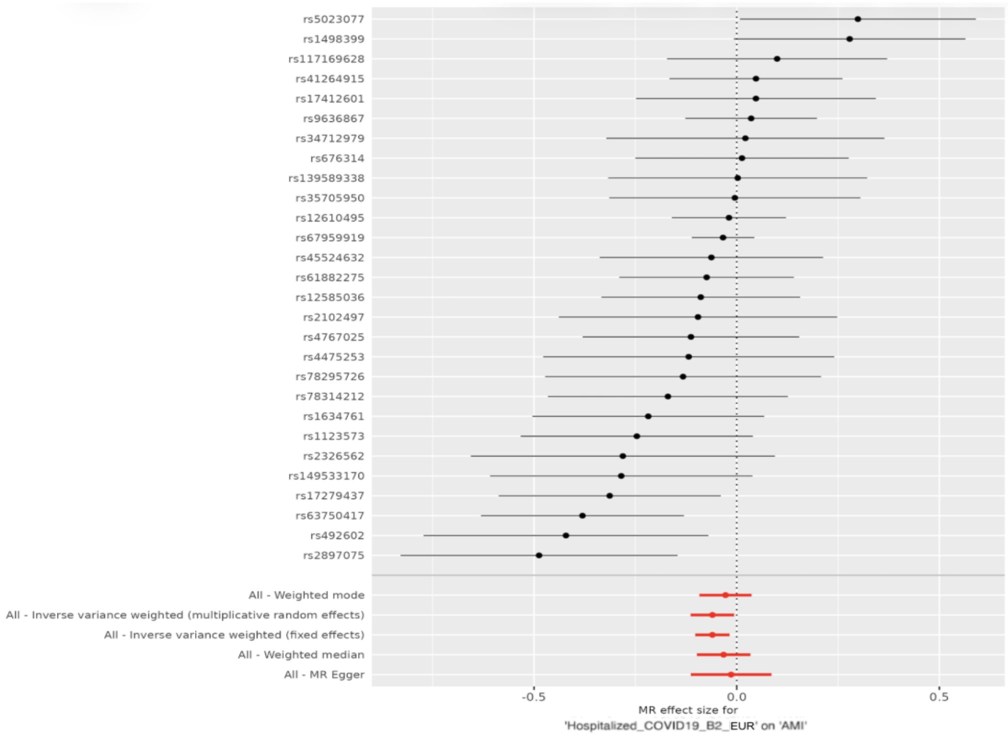


B

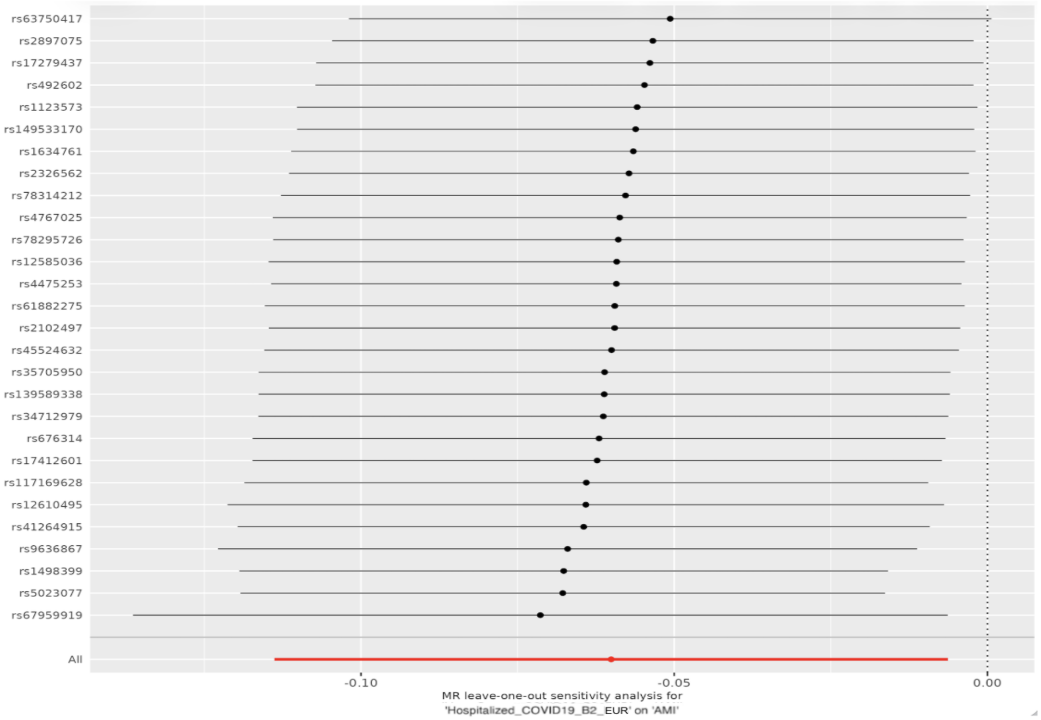

C

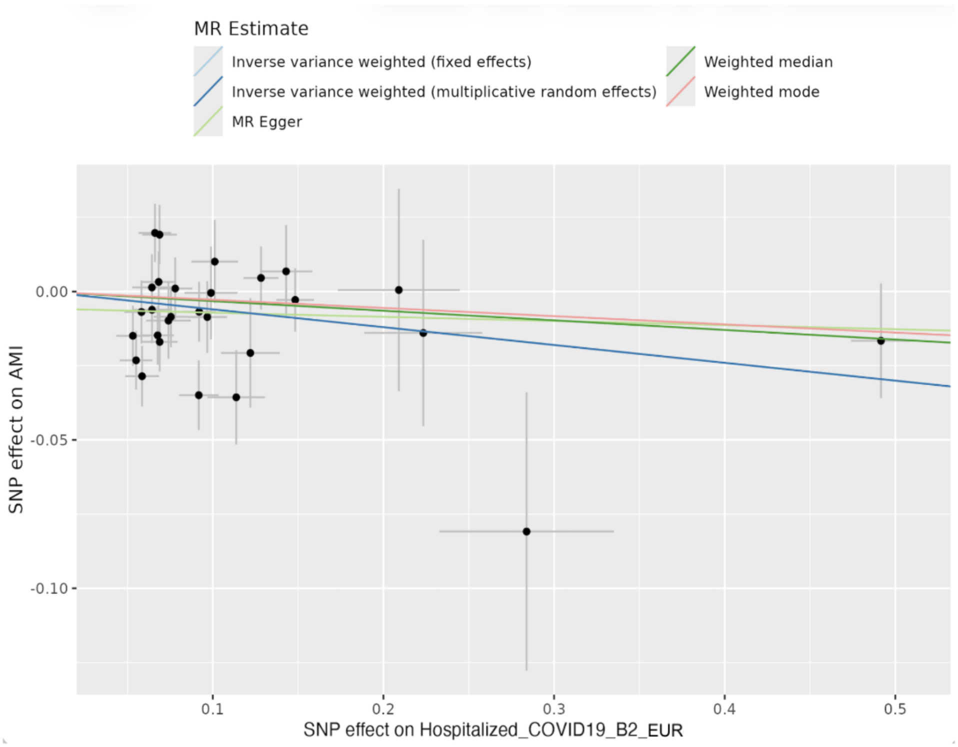


D

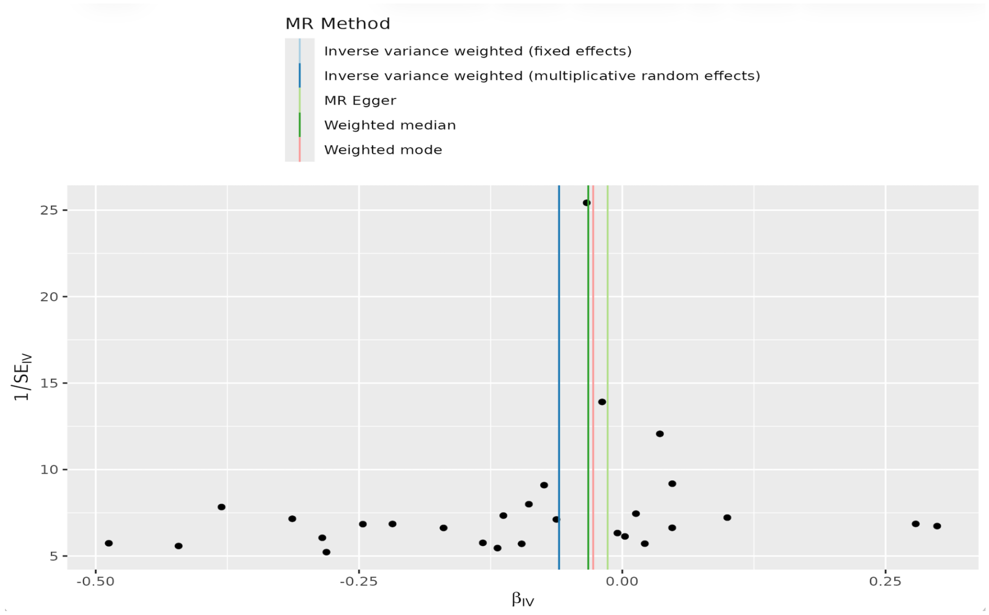


(A) Forest plot of individual SNP causal estimates. (B) Leave-one-out sensitivity analysis. (C) Scatter plot of SNP effects on COVID-19 and AMI with MR regression lines. (D) Funnel plot assessing instrument heterogeneity and potential pleiotropy. Abbreviations: AMI, acute myocardial infarction; MR, Mendelian randomization.

**Supplementary Figure 3.** MR analyses evaluating the causal association between very severe COVID-19 and AMI in European ancestry.

A

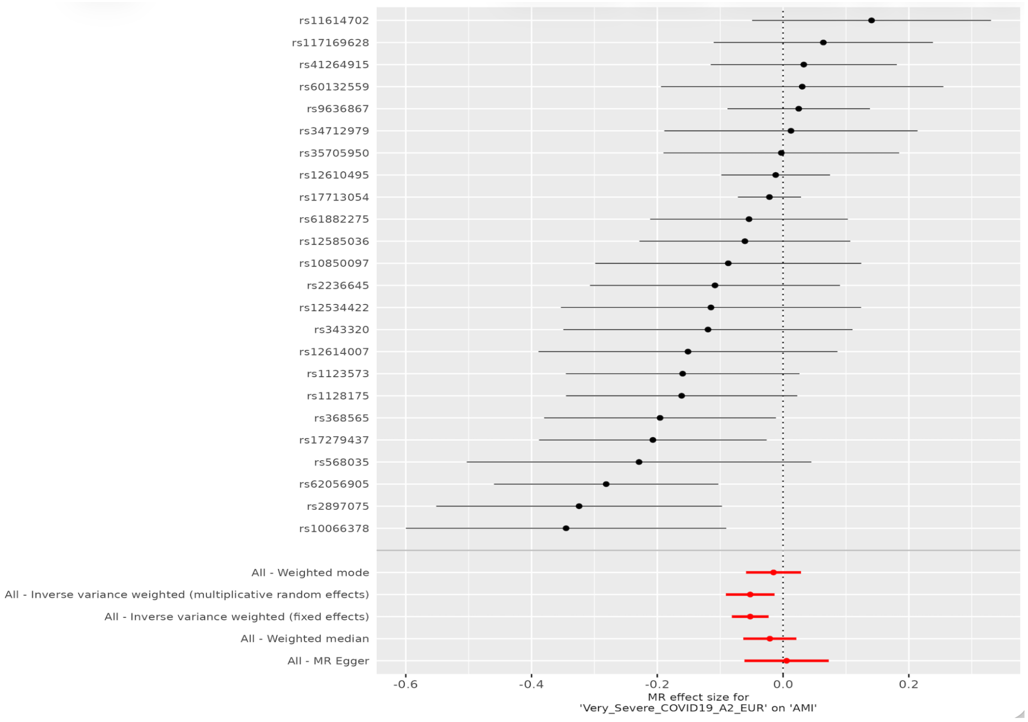


B

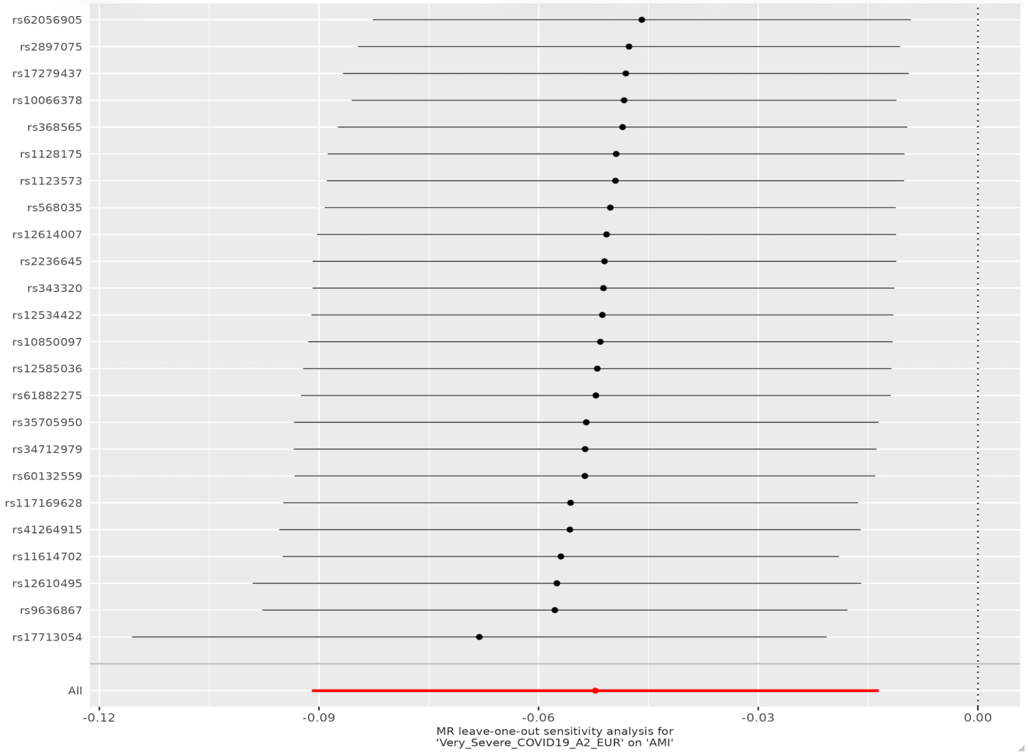


C

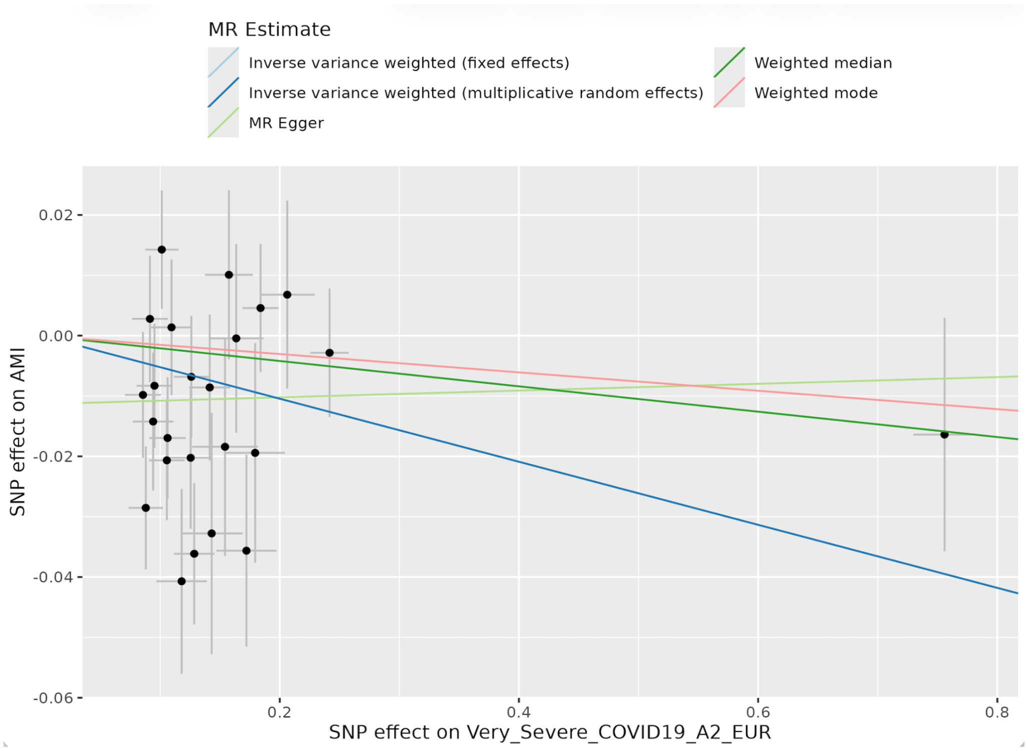


D

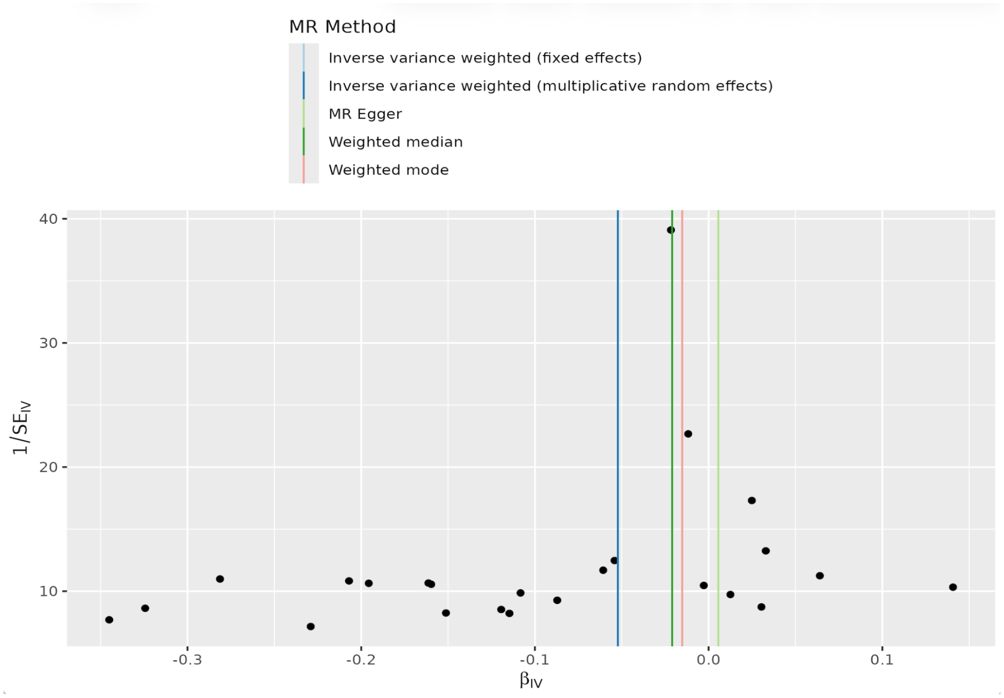


(A) Forest plot of individual SNP causal estimates. (B) Leave-one-out sensitivity analysis. (C) Scatter plot of SNP effects on COVID-19 and AMI with MR regression lines. (D) Funnel plot assessing instrument heterogeneity and potential pleiotropy. Abbreviations: AMI, acute myocardial infarction; MR, Mendelian randomization.

**Supplementary Figure 4.** MR analyses evaluating the causal association between COVID-19 infection and AMI in East Asian ancestry.

A

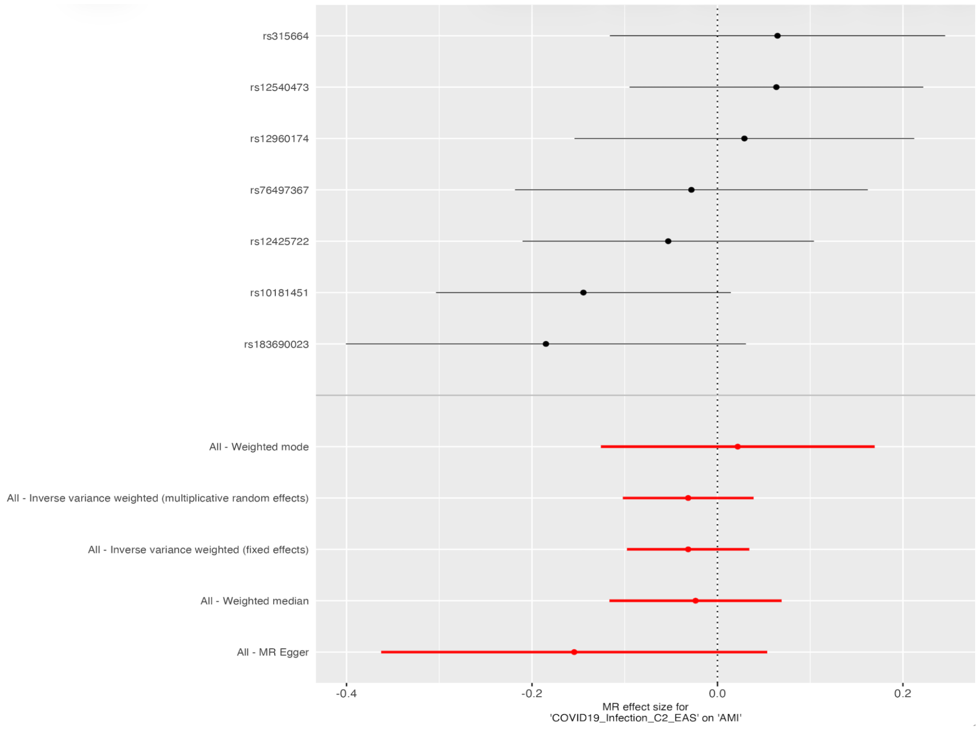


B

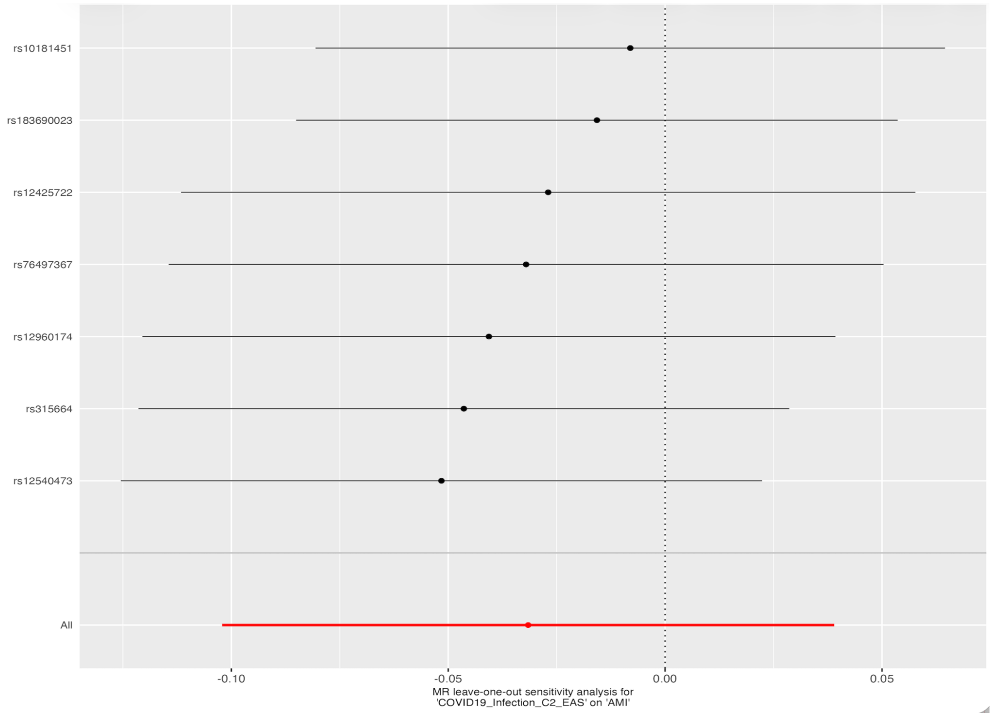

C

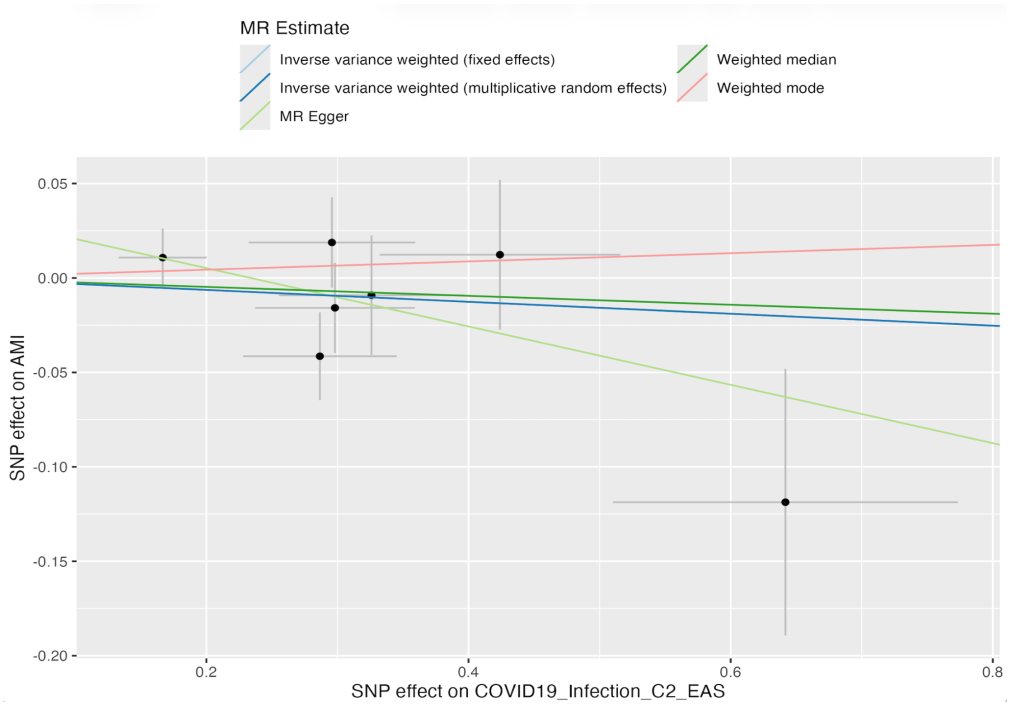


D

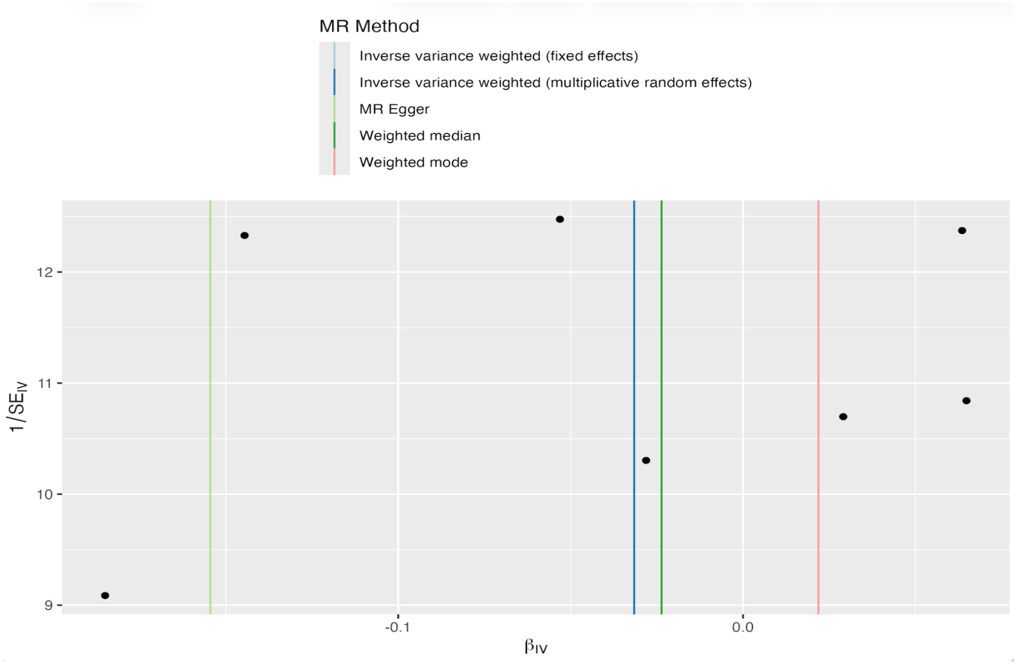


(A) Forest plot of individual SNP causal estimates. (B) Leave-one-out sensitivity analysis. (C) Scatter plot of SNP effects on COVID-19 and AMI with MR regression lines. (D) Funnel plot assessing instrument heterogeneity and potential pleiotropy. Abbreviations: AMI, acute myocardial infarction; MR, Mendelian randomization.

**Supplementary Figure 5.** MR analyses evaluating the causal association between hospitalized COVID-19 and AMI in East Asian ancestry.

A

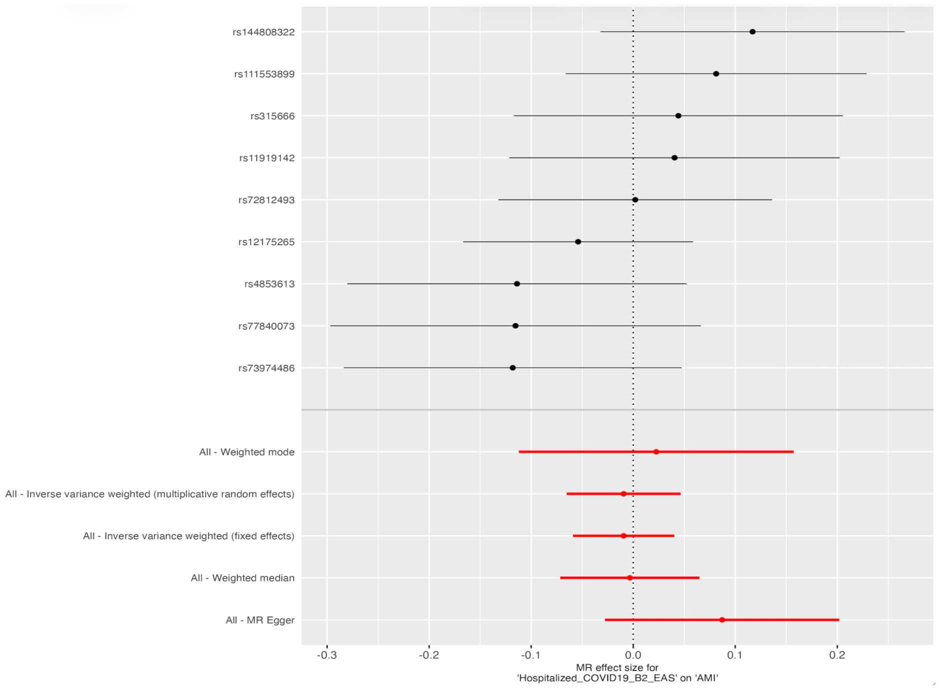


B

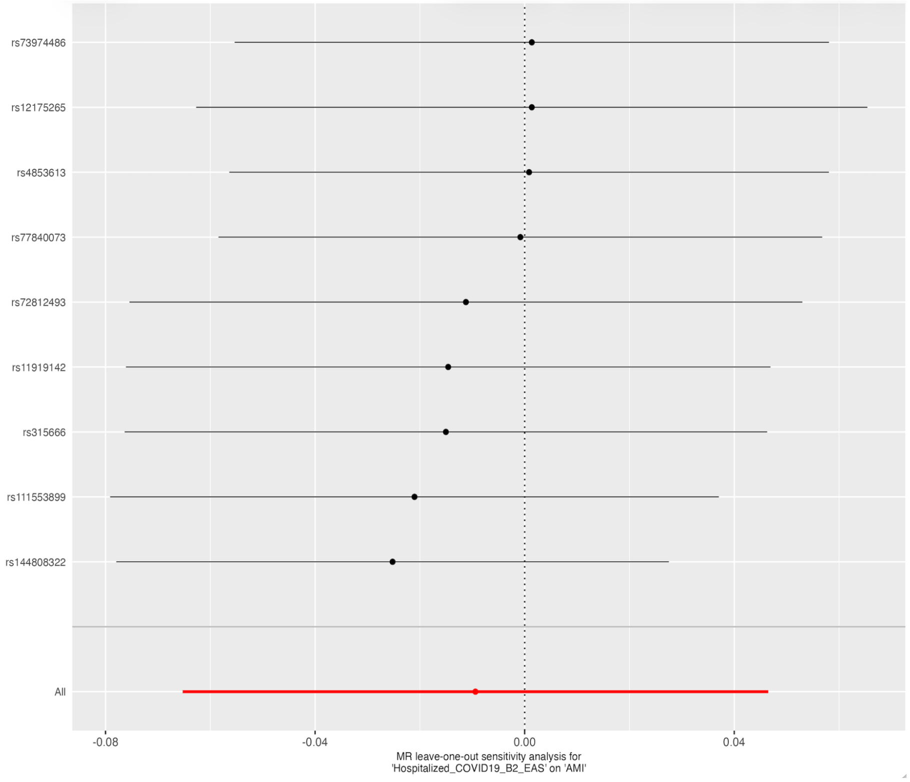

C

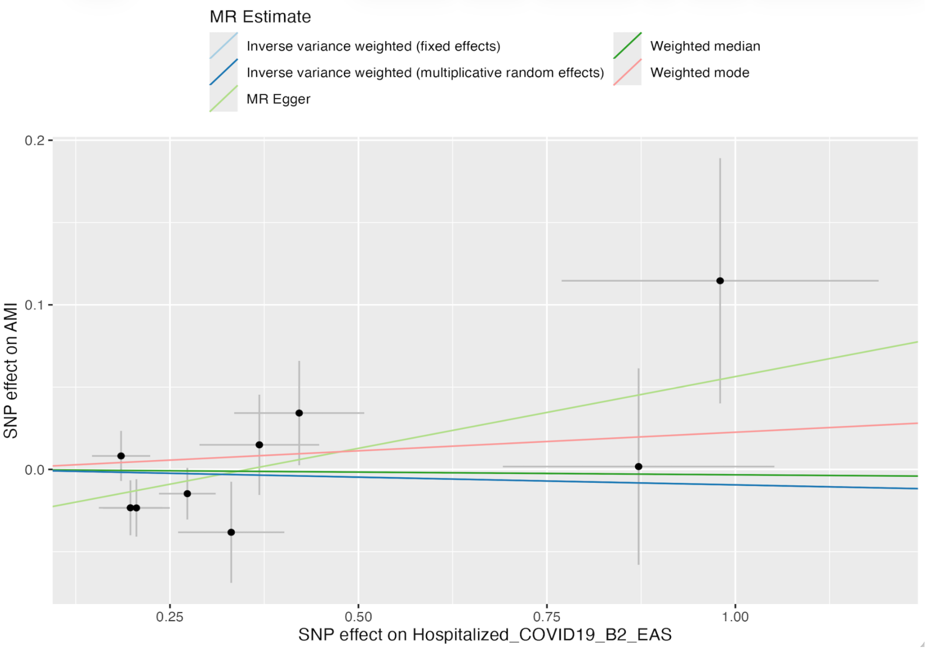


D

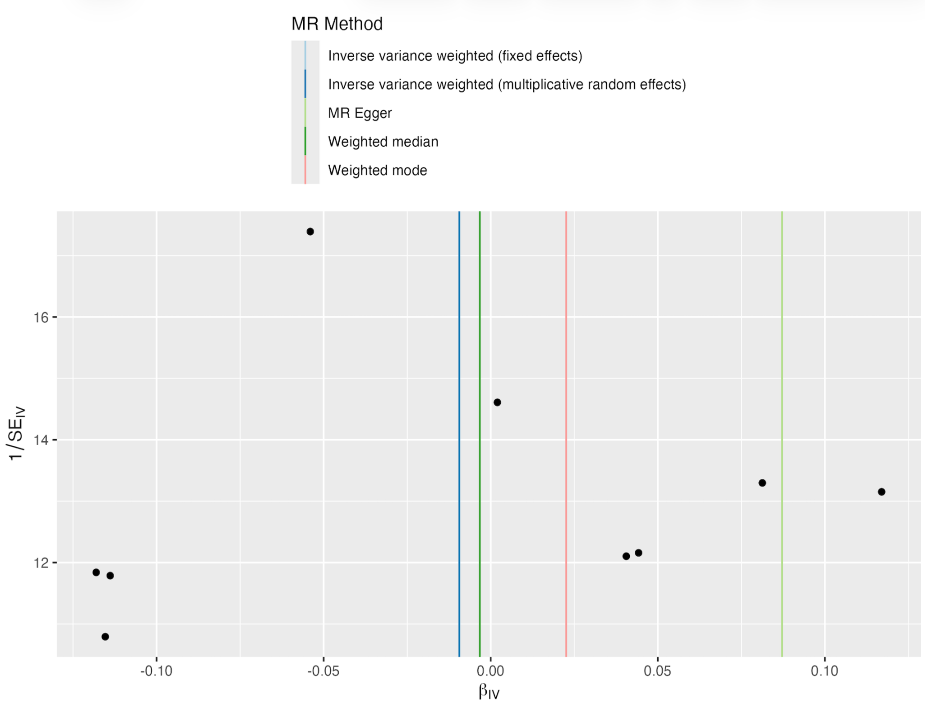


(A) Forest plot of individual SNP causal estimates. (B) Leave-one-out sensitivity analysis. (C) Scatter plot of SNP effects on COVID-19 and AMI with MR regression lines. (D) Funnel plot assessing instrument heterogeneity and potential pleiotropy. Abbreviations: AMI, acute myocardial infarction; MR, Mendelian randomization.

**Supplementary Figure 6.** MR analyses evaluating the causal association between very severe COVID-19 and AMI in East Asian ancestry.

A

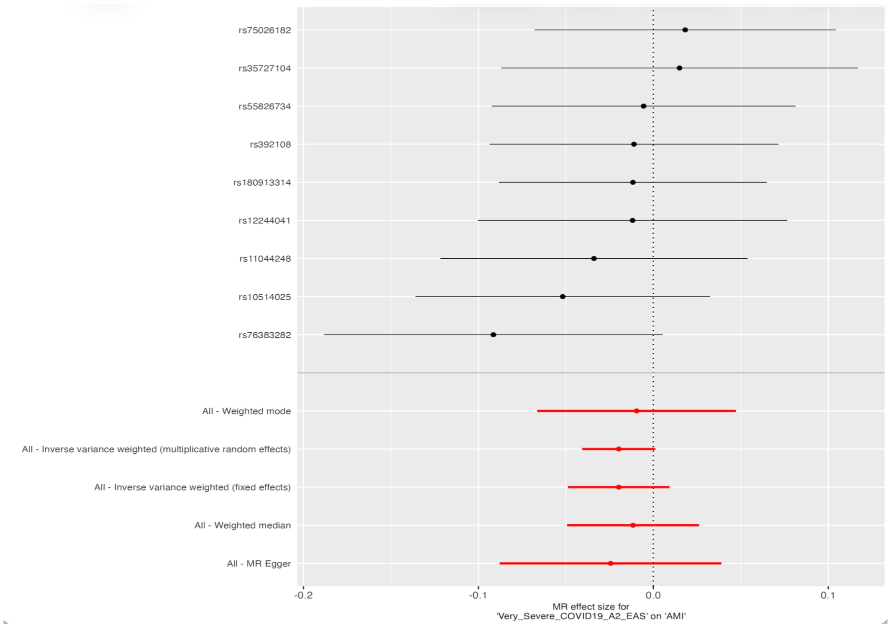


B

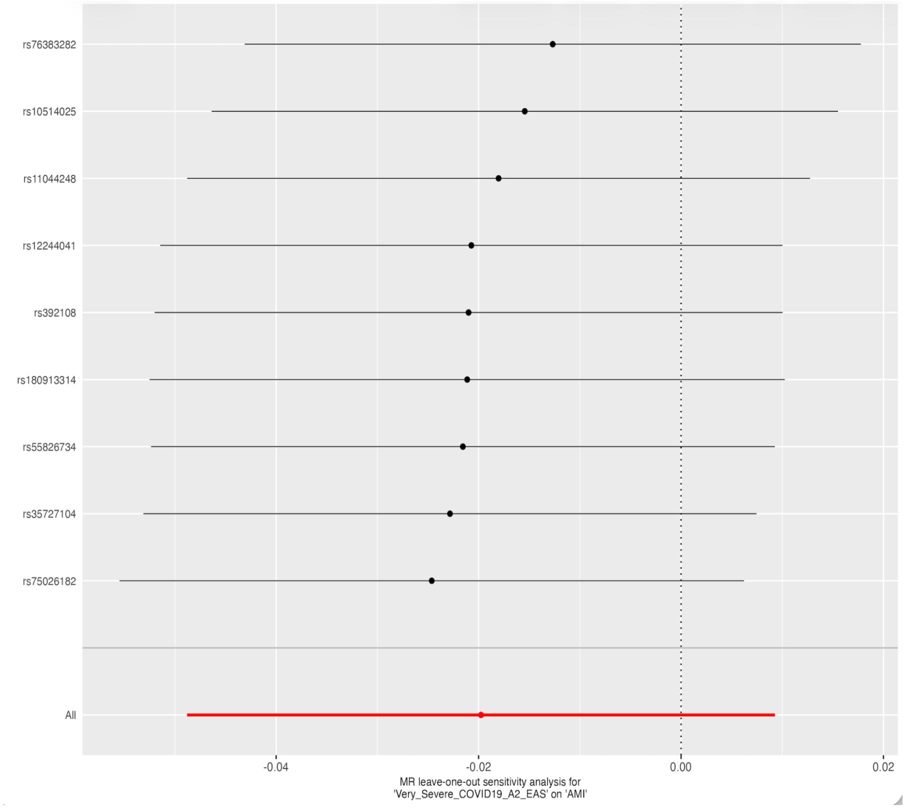

C

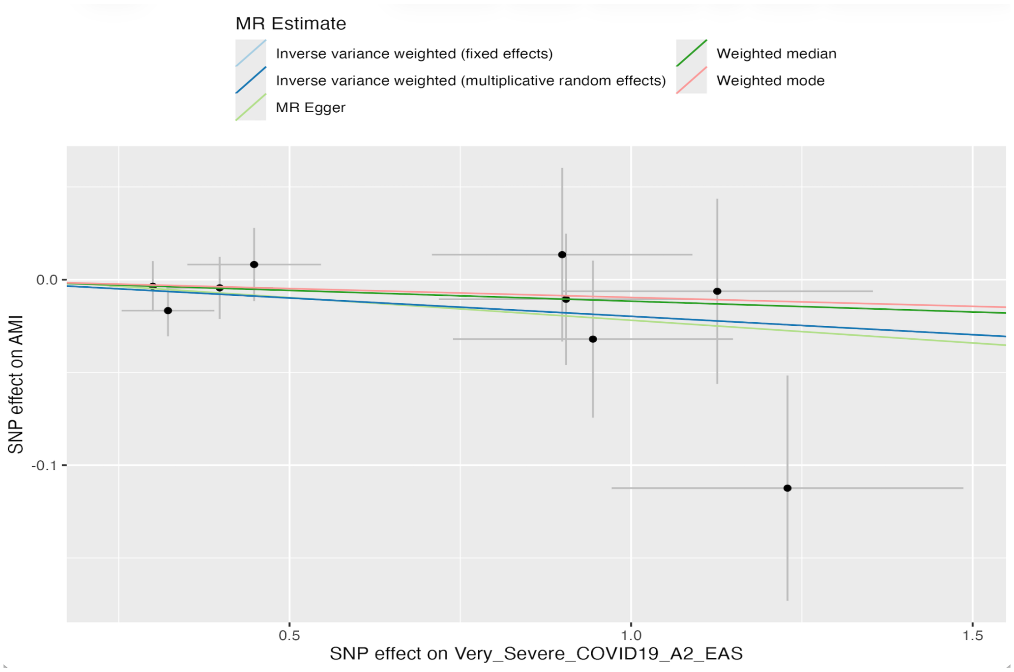


D

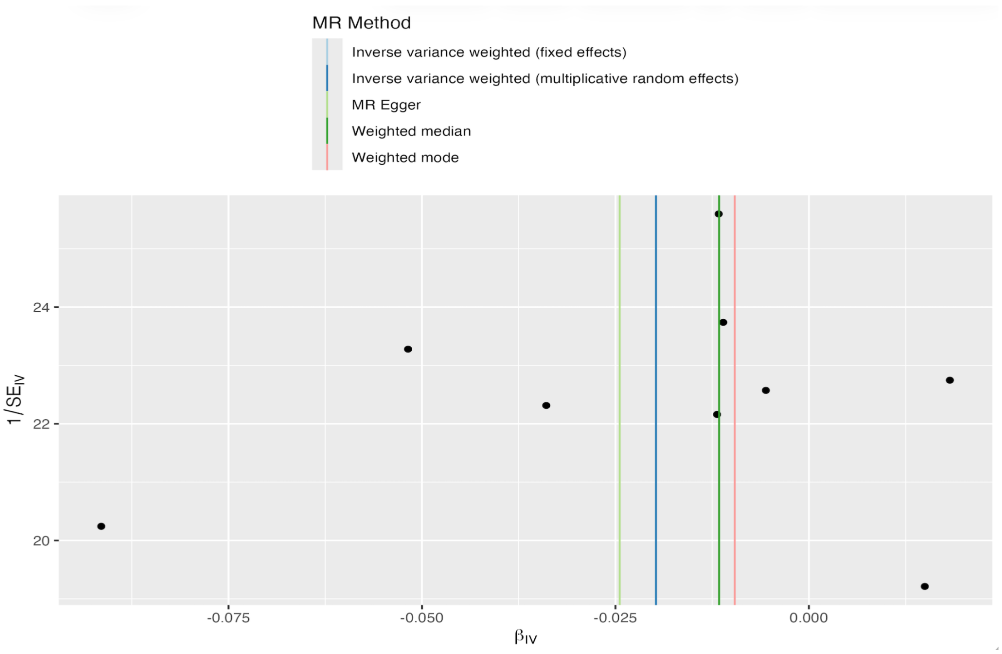


(A) Forest plot of individual SNP causal estimates. (B) Leave-one-out sensitivity analysis. (C) Scatter plot of SNP effects on COVID-19 and AMI with MR regression lines. (D) Funnel plot assessing instrument heterogeneity and potential pleiotropy. Abbreviations: AMI, acute myocardial infarction; MR, Mendelian randomization.

**Supplementary Figure 7.** MR analyses evaluating the causal association between COVID-19 infection and AMI in South Asian ancestry.

A

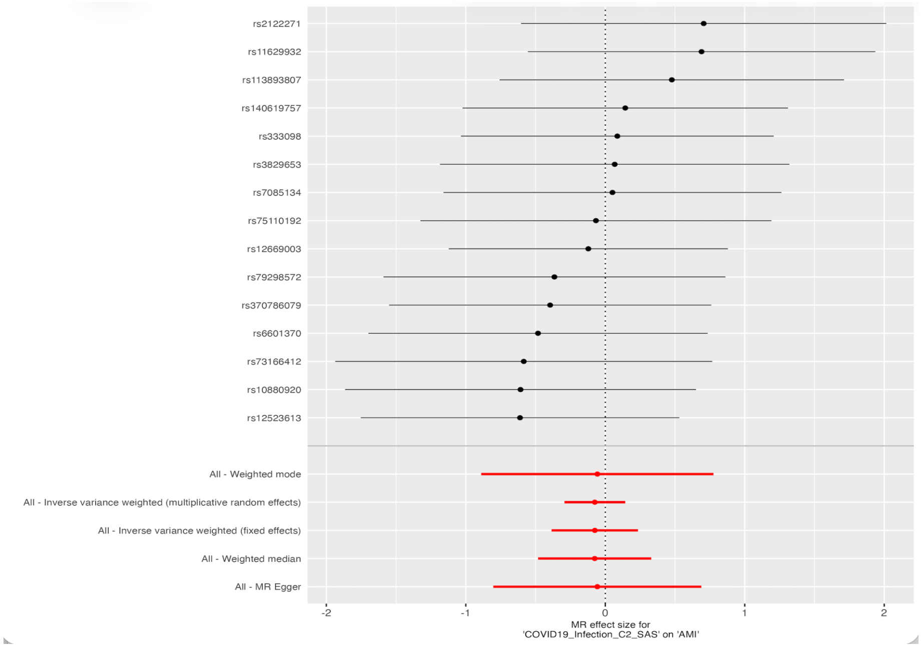


B

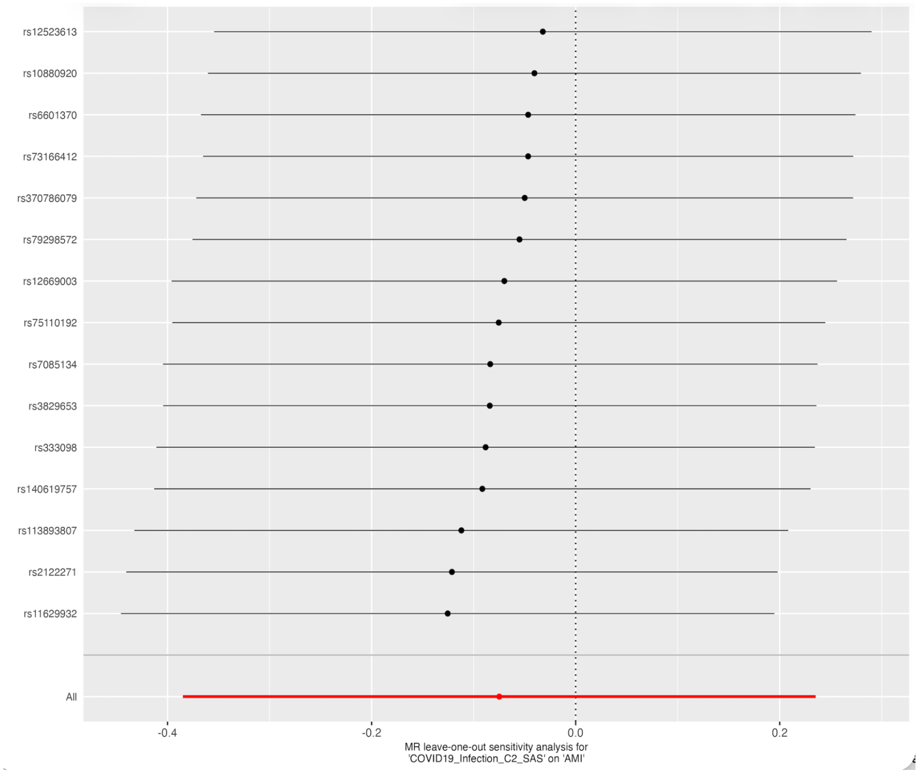

C

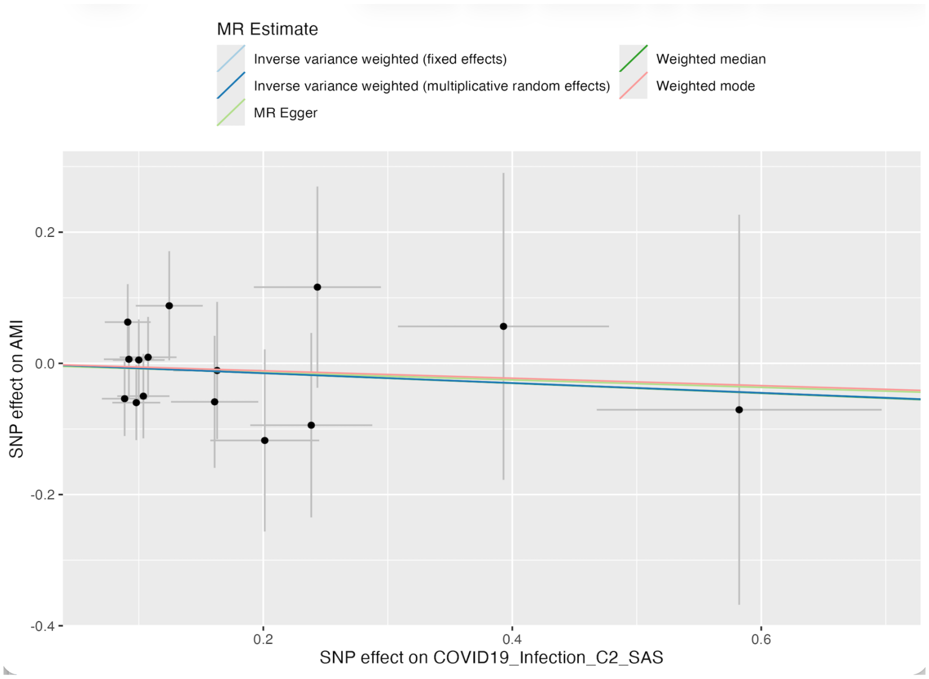


D

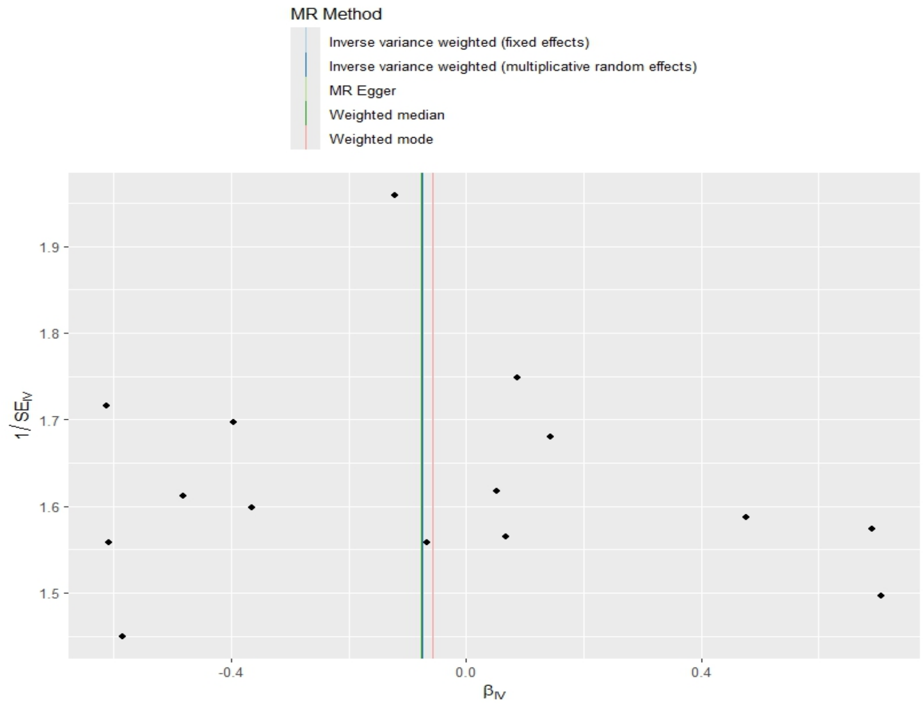


(A) Forest plot of individual SNP causal estimates. (B) Leave-one-out sensitivity analysis. (C) Scatter plot of SNP effects on COVID-19 and AMI with MR regression lines. (D) Funnel plot assessing instrument heterogeneity and potential pleiotropy. Abbreviations: AMI, acute myocardial infarction; MR, Mendelian randomization.

**Supplementary Figure 8.** MR analyses evaluating the causal association between hospitalized COVID-19 and AMI in South Asian ancestry.

A

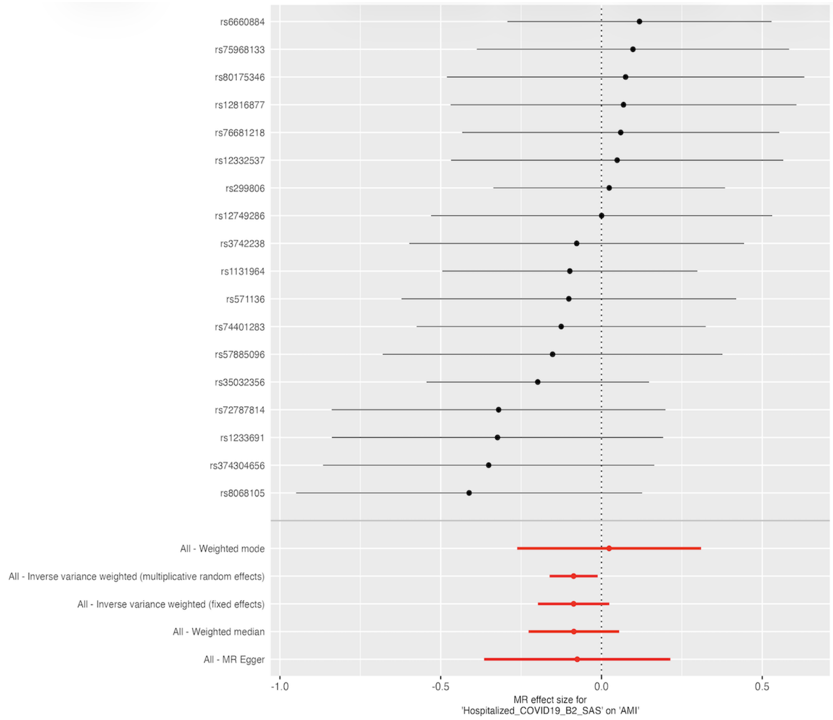


B

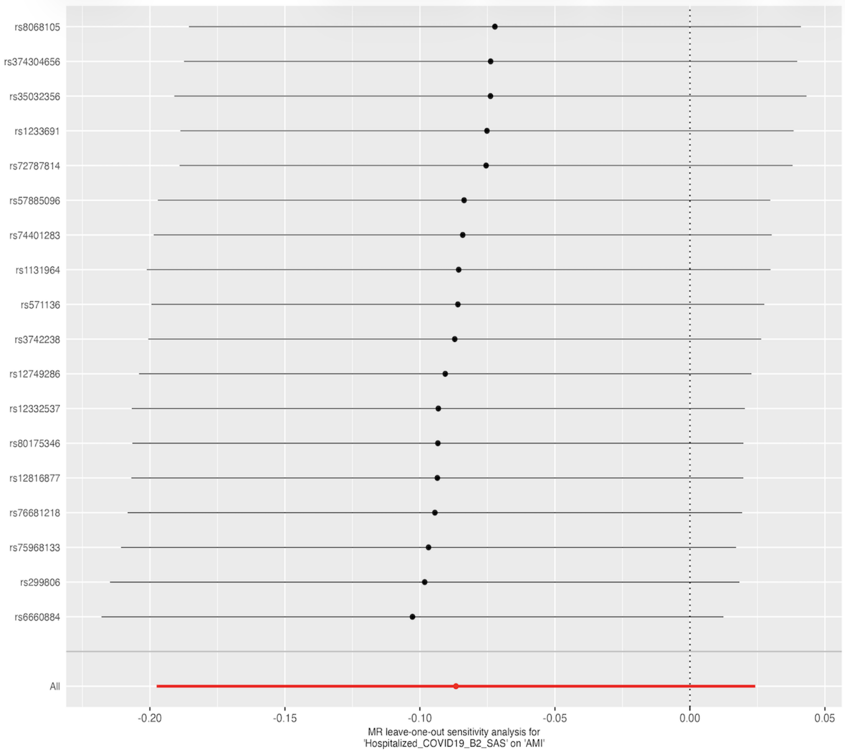

C

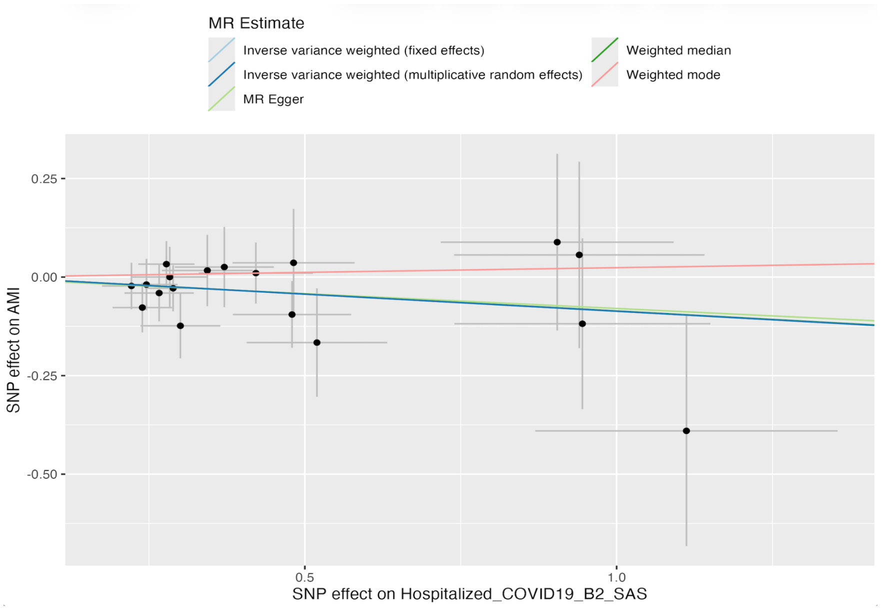


D

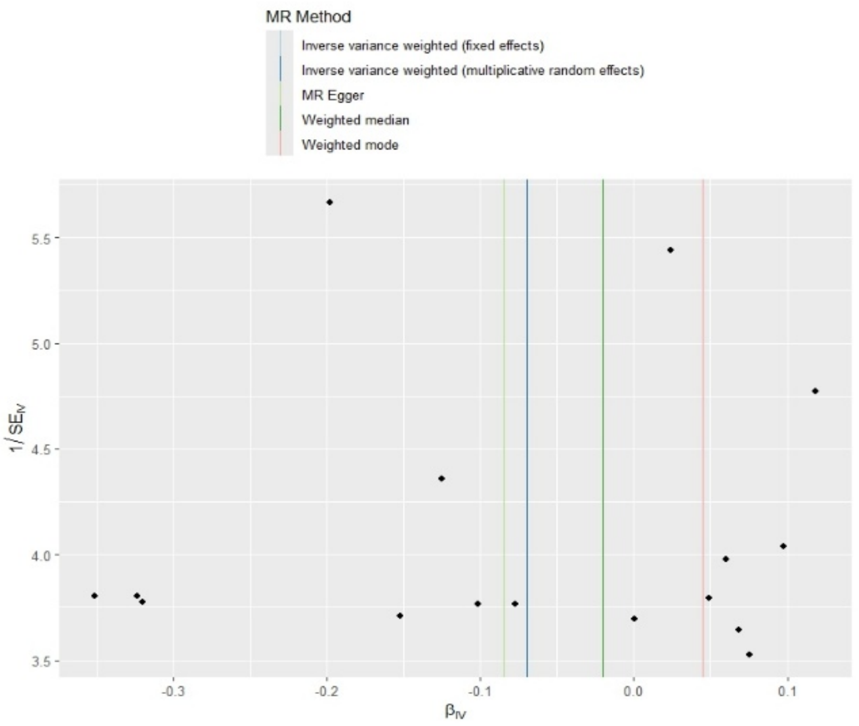


(A) Forest plot of individual SNP causal estimates. (B) Leave-one-out sensitivity analysis. (C) Scatter plot of SNP effects on COVID-19 and AMI with MR regression lines. (D) Funnel plot assessing instrument heterogeneity and potential pleiotropy. Abbreviations: AMI, acute myocardial infarction; MR, Mendelian randomization.

**Supplementary Figure 9.** MR analyses evaluating the causal association between very severe COVID-19 and AMI in South Asian ancestry.

A

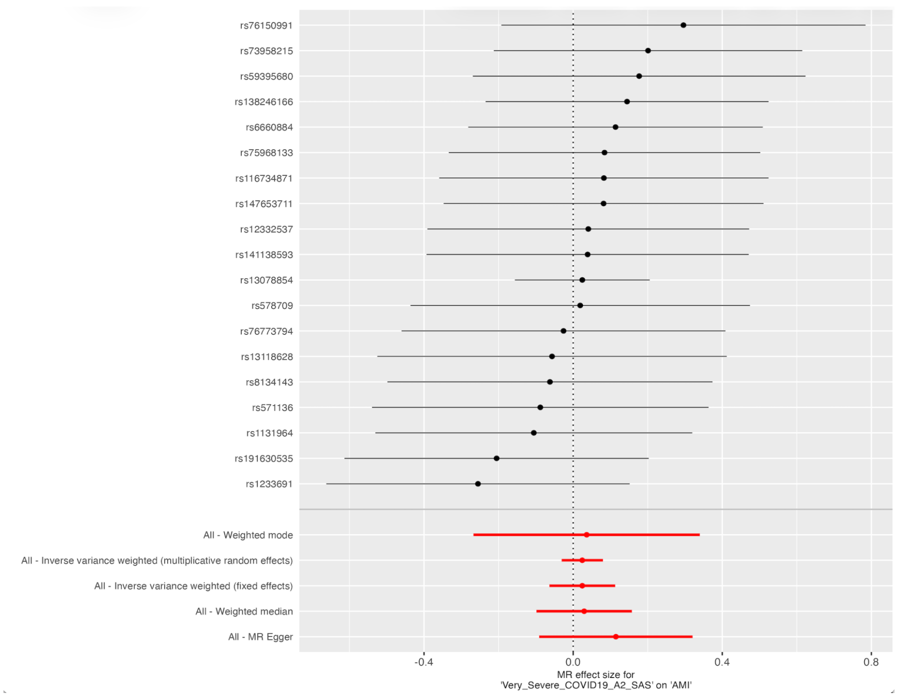


B

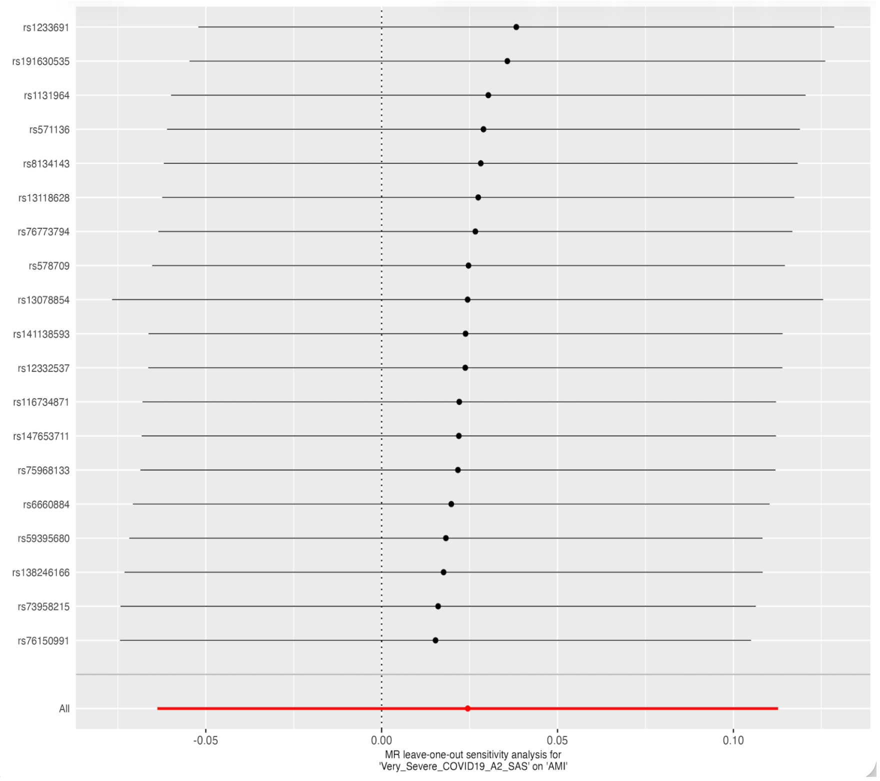

C

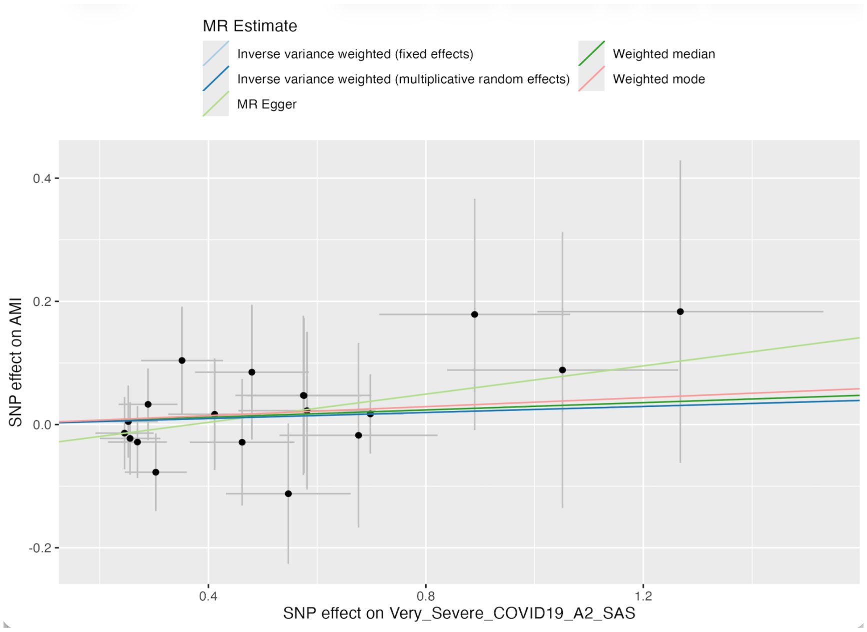


D

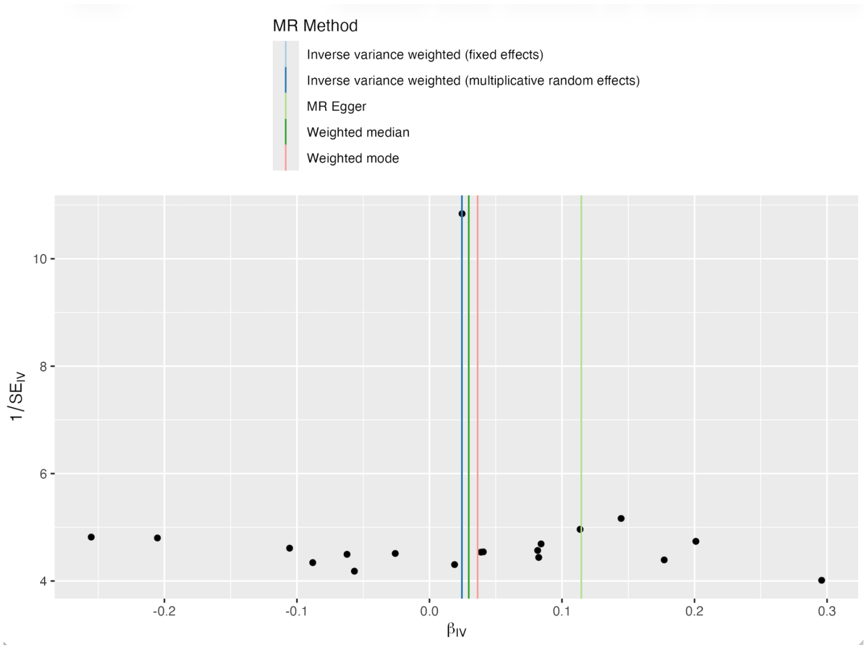


(A) Forest plot of individual SNP causal estimates. (B) Leave-one-out sensitivity analysis. (C) Scatter plot of SNP effects on COVID-19 and AMI with MR regression lines. (D) Funnel plot assessing instrument heterogeneity and potential pleiotropy. Abbreviations: AMI, acute myocardial infarction; MR, Mendelian randomization.

**Supplementary Figure 10.** MR analyses evaluating the causal association between COVID-19 infection and AMI in African ancestry.

A

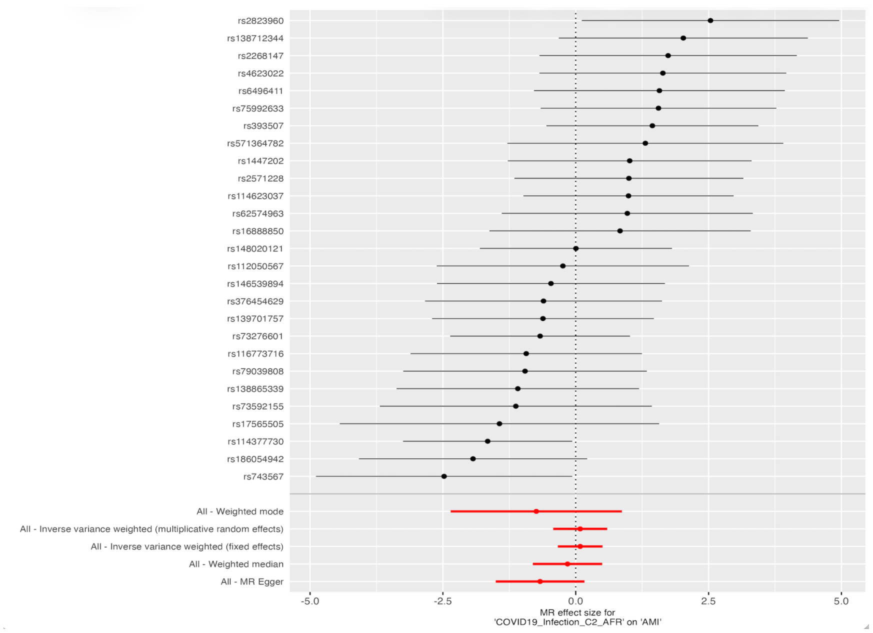


B

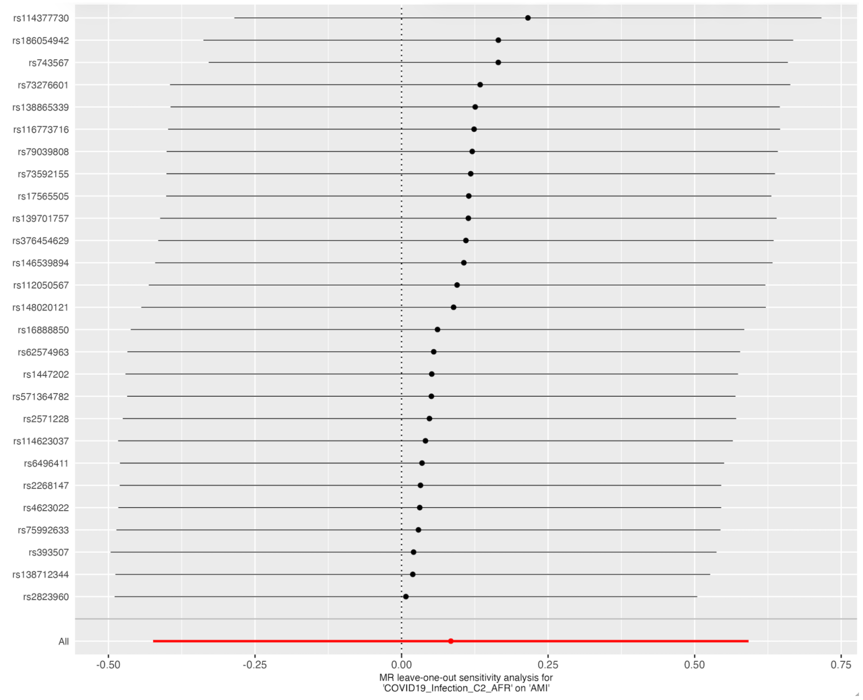

C

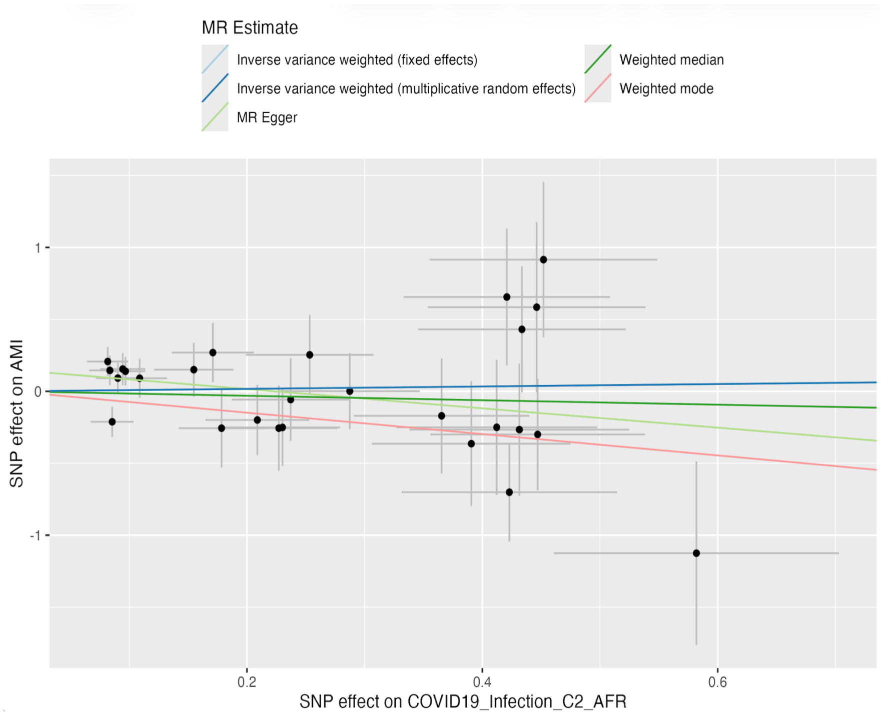


D

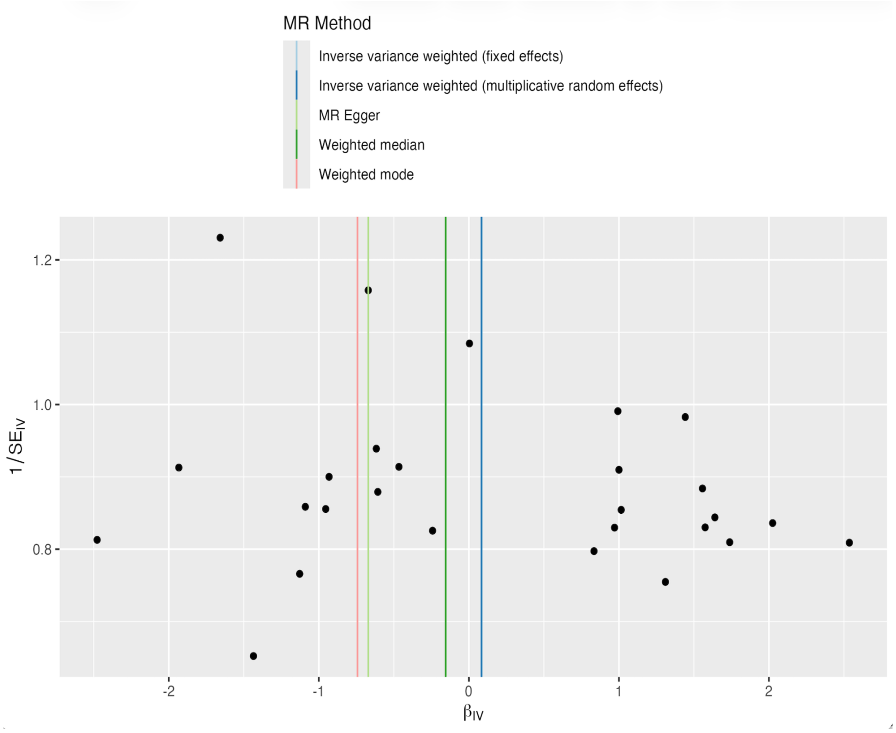


(A) Forest plot of individual SNP causal estimates. (B) Leave-one-out sensitivity analysis. (C) Scatter plot of SNP effects on COVID-19 and AMI with MR regression lines. (D) Funnel plot assessing instrument heterogeneity and potential pleiotropy. Abbreviations: AMI, acute myocardial infarction; MR, Mendelian randomization.

**Supplementary Figure 11.** MR analyses evaluating the causal association between hospitalized COVID-19 and AMI in African ancestry.

A

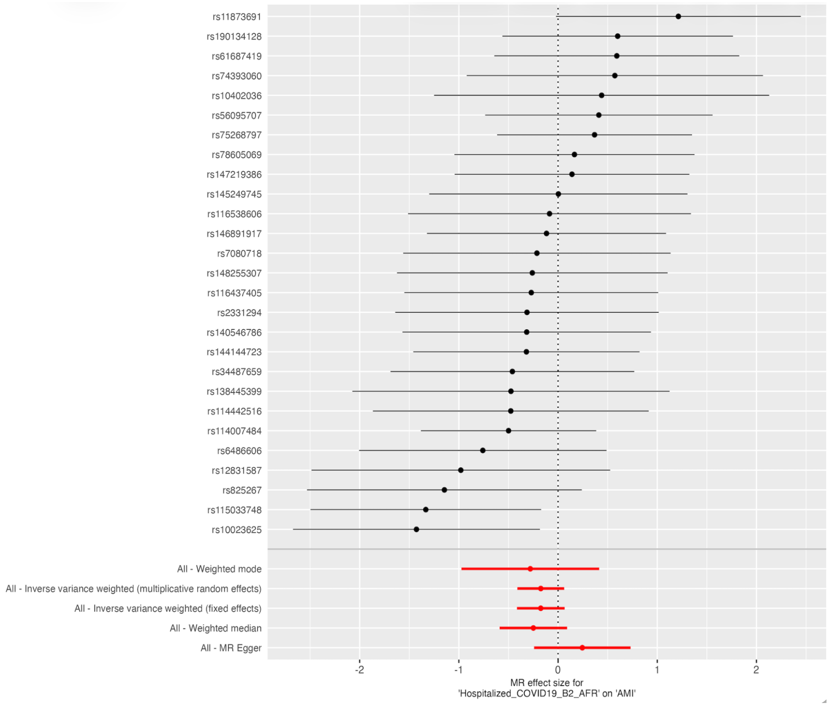


B

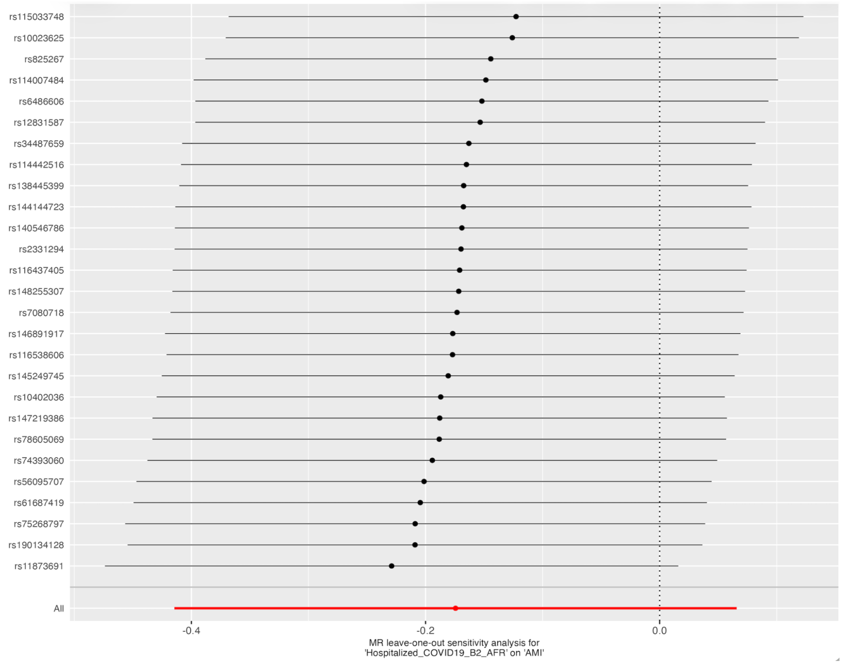

C

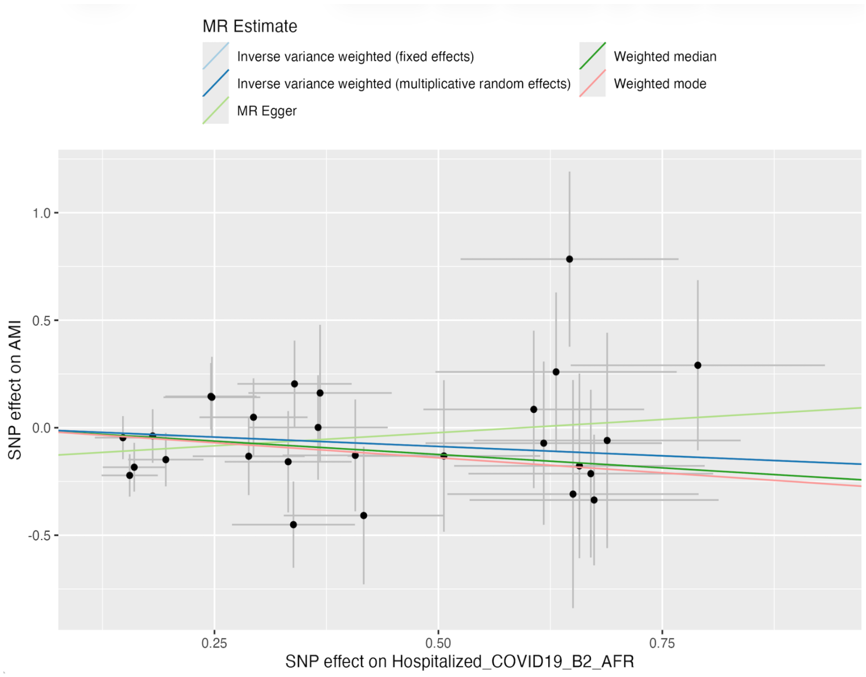


D

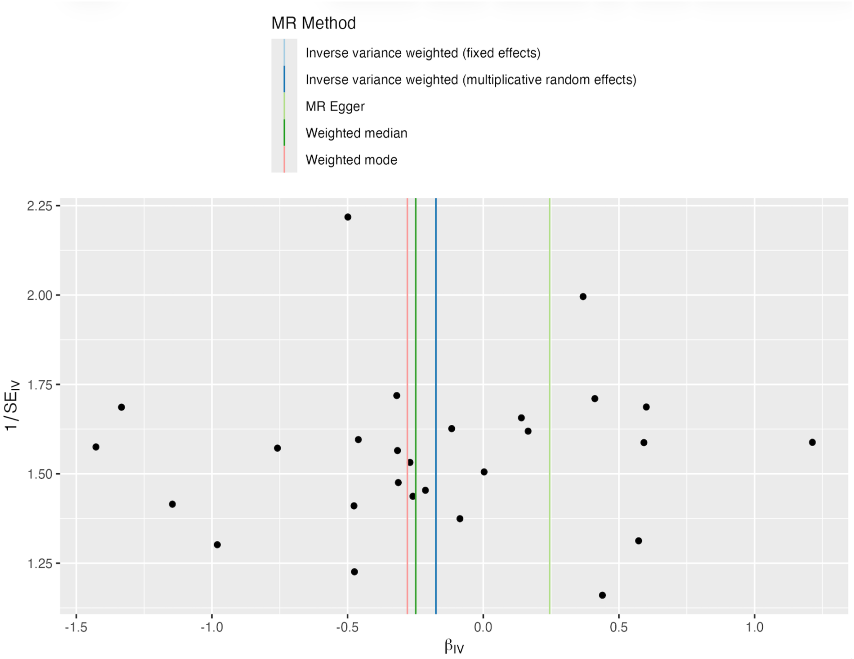


(A) Forest plot of individual SNP causal estimates. (B) Leave-one-out sensitivity analysis. (C) Scatter plot of SNP effects on COVID-19 and AMI with MR regression lines. (D) Funnel plot assessing instrument heterogeneity and potential pleiotropy. Abbreviations: AMI, acute myocardial infarction; MR, Mendelian randomization.

**Supplementary Figure 12.** MR analyses evaluating the causal association between very severe COVID-19 and AMI in African ancestry.

A

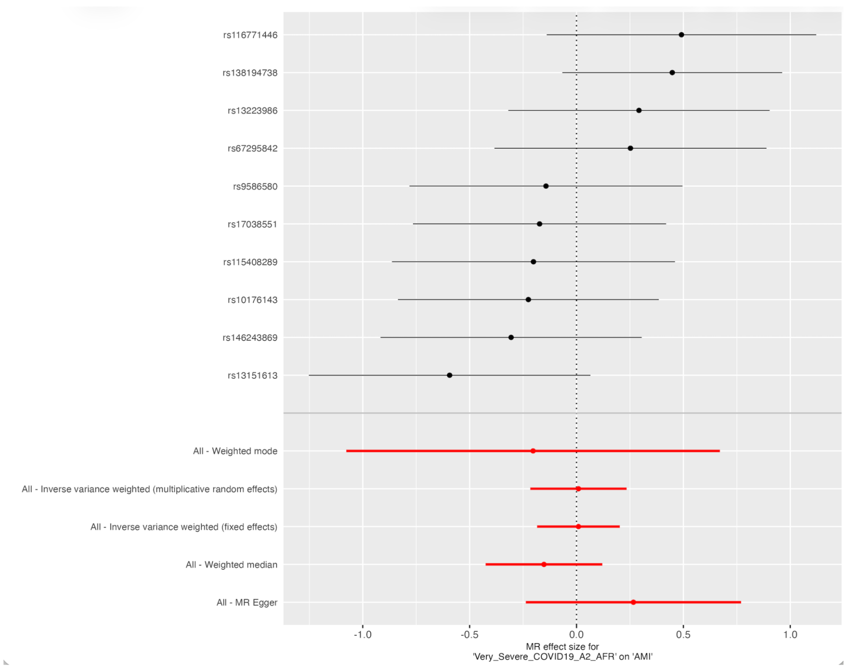


B

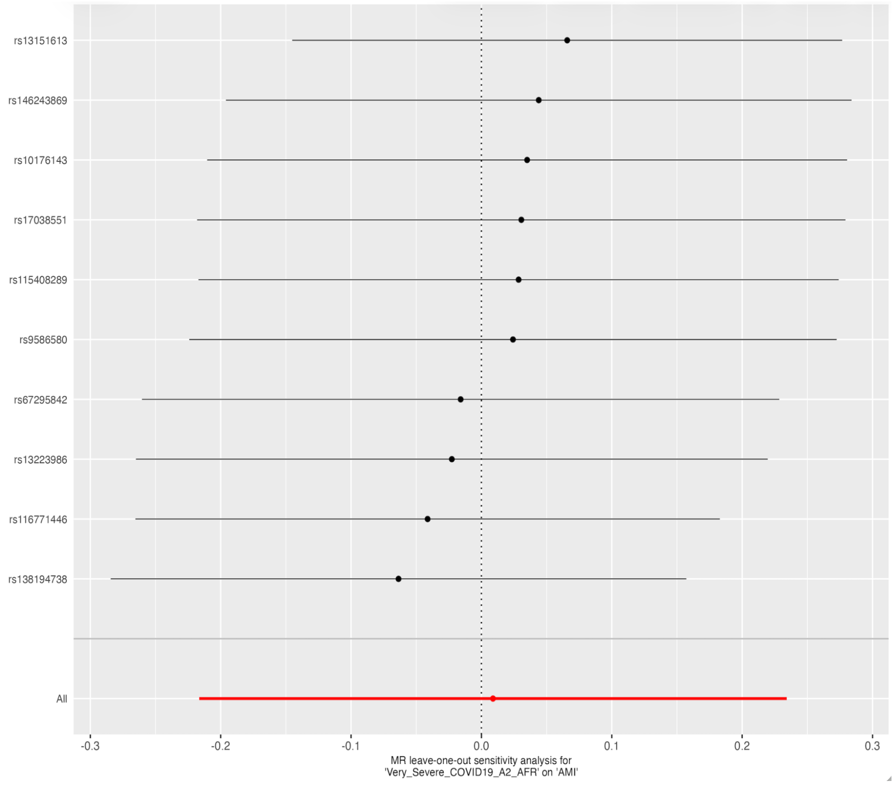


C

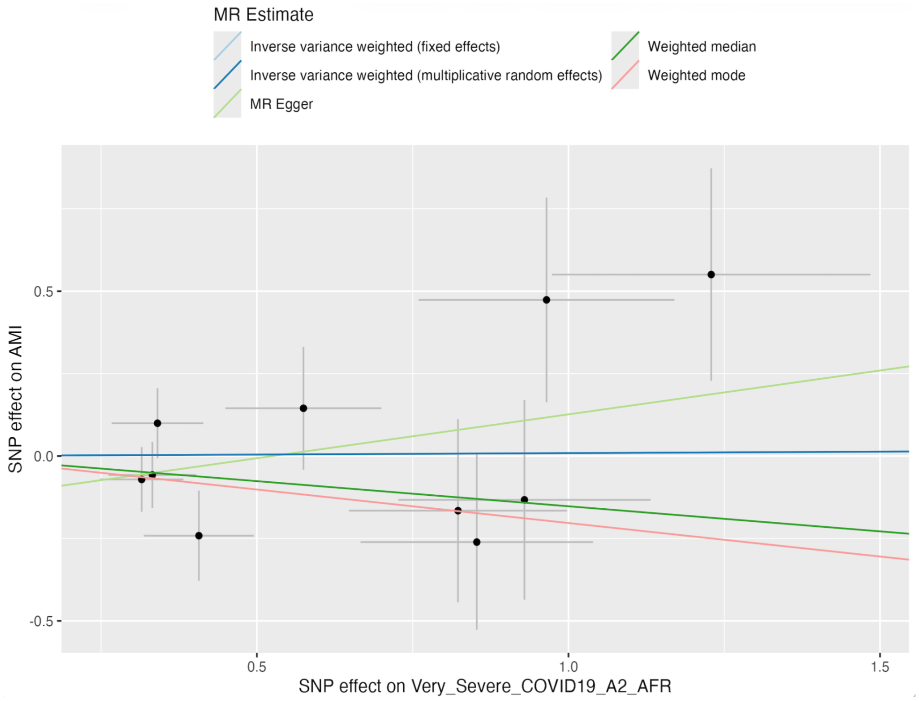


D

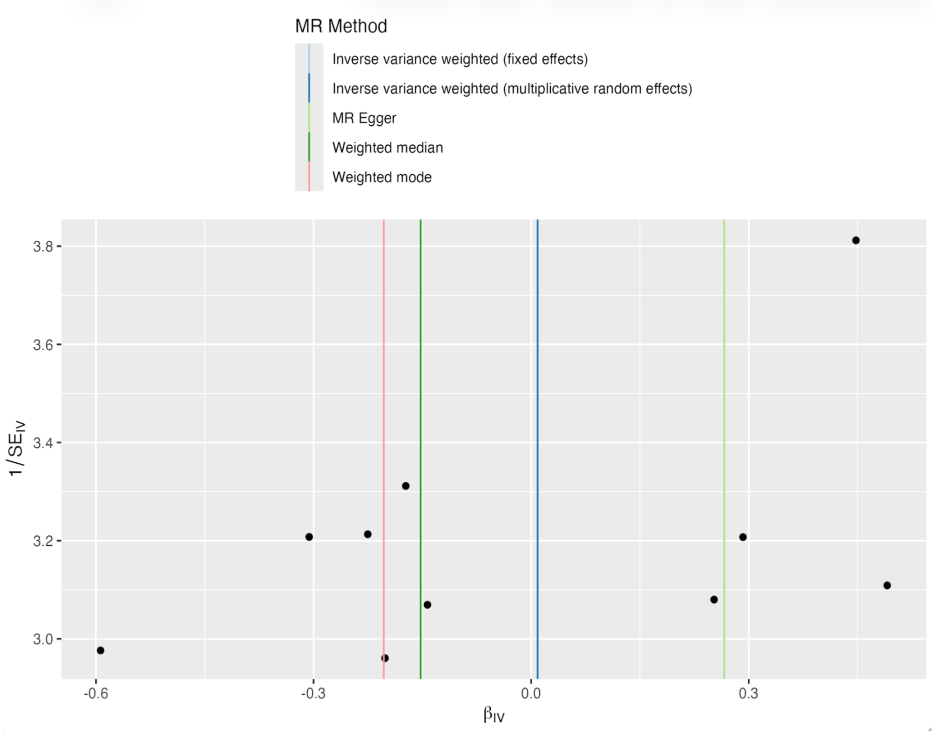


(A) Forest plot of individual SNP causal estimates. (B) Leave-one-out sensitivity analysis. (C) Scatter plot of SNP effects on COVID-19 and AMI with MR regression lines. (D) Funnel plot assessing instrument heterogeneity and potential pleiotropy. Abbreviations: AMI, acute myocardial infarction; MR, Mendelian randomization.

#### Supplementary Tables

**Supplementary Table 1.** Characteristics of genetic variants used as IVs for COVID-19 infection in the European MR analysis

| rsID | Chr | Pos (GRCh38) | EA | NEA | Beta | SE | P-value |
|---|---|---|---|---|---|---|---|
| rs10774673 | 12 | 112923353 | C | T | 0.028916 | 0.00483 | 2.14E-09 |
| rs1123573 | 2 | 60480453 | A | G | -0.02639 | 0.004769 | 3.16E-08 |
| rs11264339 | 1 | 155168172 | C | T | -0.03502 | 0.004562 | 1.62E-14 |
| rs12610495 | 19 | 4717660 | A | G | 0.049515 | 0.005077 | 1.80E-22 |
| rs184781326 | 14 | 28990028 | A | G | -0.06902 | 0.011679 | 3.42E-09 |
| rs2260685 | 3 | 195770872 | T | C | 0.033425 | 0.004689 | 1.01E-12 |
| rs2290859 | 3 | 101806781 | C | T | -0.05084 | 0.00486 | 1.32E-25 |
| rs2834158 | 21 | 33244908 | T | C | -0.04055 | 0.004871 | 8.51E-17 |
| rs35044562 | 3 | 45867532 | A | G | 0.12798 | 0.008476 | 1.64E-51 |
| rs676314 | 19 | 50362278 | A | G | 0.027577 | 0.004815 | 1.02E-08 |
| rs7118388 | 11 | 34432600 | A | G | 0.026551 | 0.004533 | 4.71E-09 |
| rs73062389 | 3 | 45793925 | G | A | 0.20044 | 0.009861 | 7.61E-92 |
| rs9264740 | 6 | 31276554 | C | T | -0.03047 | 0.00526 | 6.87E-09 |

**Footnote:** IVs - instrumental variables; rsID - variants ID; Chr - Chromosome; Pos - base pair location; EA - effect allele; NEA - non-effect allele; SE - standard error.

**Supplementary Table 2.** Characteristics of genetic variants used as IVs for hospitalized COVID-19 in the European MR analysis

| rsID | Chr | Pos (GRCh38) | EA | NEA | Beta | SE | P-value |
|---|---|---|---|---|---|---|---|
| rs1123573 | 2 | 60480453 | A | G | -0.06885 | 0.010432 | 4.13E-11 |
| rs117169628 | 16 | 89196249 | G | A | 0.10114 | 0.013651 | 1.27E-13 |
| rs12585036 | 13 | 112881427 | C | T | 0.096736 | 0.011638 | 9.39E-17 |
| rs12610495 | 19 | 4717660 | A | G | 0.14828 | 0.011043 | 4.21E-41 |
| rs139589338 | 1 | 154853813 | A | G | 0.20902 | 0.03574 | 4.97E-09 |
| rs149533170 | 9 | 21172826 | G | A | 0.28389 | 0.051106 | 2.78E-08 |
| rs1498399 | 1 | 77481554 | A | G | 0.068773 | 0.010063 | 8.24E-12 |
| rs1634761 | 6 | 31306250 | C | T | -0.06768 | 0.009495 | 1.02E-12 |
| rs17279437 | 3 | 45772602 | G | A | -0.11372 | 0.016819 | 1.37E-11 |
| rs17412601 | 3 | 101780431 | T | C | -0.06826 | 0.010317 | 3.69E-11 |
| rs2102497 | 16 | 54221310 | T | C | 0.064371 | 0.011743 | 4.21E-08 |
| rs2326562 | 8 | 60515641 | C | T | 0.053141 | 0.009736 | 4.81E-08 |
| rs2897075 | 7 | 100032719 | C | T | 0.058531 | 0.009857 | 2.88E-09 |
| rs34712979 | 4 | 105897896 | G | A | -0.06431 | 0.011488 | 2.17E-08 |
| rs35705950 | 11 | 1219991 | G | T | -0.09893 | 0.015532 | 1.90E-10 |
| rs41264915 | 1 | 155197995 | A | G | -0.14296 | 0.015375 | 1.43E-20 |
| rs4475253 | 5 | 132440814 | A | G | 0.058194 | 0.010052 | 7.07E-09 |
| rs45524632 | 19 | 10486312 | C | A | 0.22336 | 0.034592 | 1.07E-10 |
| rs4767025 | 12 | 112920989 | C | T | 0.07545 | 0.010079 | 7.13E-14 |
| rs492602 | 19 | 48703160 | A | G | -0.05504 | 0.009534 | 7.81E-09 |
| rs5023077 | 12 | 132565387 | T | C | -0.06608 | 0.009515 | 3.79E-12 |
| rs61882275 | 11 | 34482745 | G | A | -0.09196 | 0.009945 | 2.30E-20 |
| rs63750417 | 17 | 45983409 | C | T | -0.09176 | 0.011487 | 1.37E-15 |
| rs676314 | 19 | 50362278 | A | G | 0.077962 | 0.010055 | 8.95E-15 |
| rs67959919 | 3 | 45830416 | G | A | 0.49158 | 0.017513 | 2.36E-173 |
| rs78295726 | 19 | 10315836 | C | T | 0.073968 | 0.013044 | 1.42E-08 |
| rs78314212 | 21 | 33940612 | C | T | 0.122 | 0.017175 | 1.22E-12 |

| | | | | | | | |
|---|---|---|---|---|---|---|---|
| rs9636867 | 21 | 33237639 | A | G | 0.12824 | 0.010176 | 2.05E-36 |

**Footnote:** IVs - instrumental variables; rsID - variants ID; Chr - Chromosome; Pos - base pair location; EA - effect allele; NEA - non-effect allele; SE - standard error.

**Supplementary Table 3.** Characteristics of genetic variants used as IVs for very severe COVID-19 in the European MR analysis

| rsID | Chr | Pos (GRCh38) | EA | NEA | Beta | SE | P-value |
|---|---|---|---|---|---|---|---|
| rs10066378 | 5 | 132441275 | T | C | 0.11796 | 0.021006 | 1.96E-08 |
| rs10850097 | 12 | 112923312 | C | T | 0.095247 | 0.014916 | 1.71E-10 |
| rs1123573 | 2 | 60480453 | A | G | -0.10621 | 0.0152 | 2.80E-12 |
| rs1128175 | 6 | 31182658 | A | G | -0.1255 | 0.017115 | 2.25E-13 |
| rs11614702 | 12 | 132481571 | G | A | 0.10135 | 0.01383 | 2.33E-13 |
| rs117169628 | 16 | 89196249 | G | A | 0.15749 | 0.020078 | 4.36E-15 |
| rs12534422 | 7 | 75634474 | C | T | 0.085624 | 0.015073 | 1.34E-08 |
| rs12585036 | 13 | 112881427 | C | T | 0.14143 | 0.017239 | 2.32E-16 |
| rs12610495 | 19 | 4717660 | A | G | 0.24167 | 0.016049 | 3.05E-51 |
| rs12614007 | 2 | 57089368 | G | A | 0.094133 | 0.016881 | 2.46E-08 |
| rs17279437 | 3 | 45772602 | G | A | -0.17214 | 0.025124 | 7.29E-12 |
| rs17713054 | 3 | 45818159 | G | A | 0.75584 | 0.026008 | 1.09E-185 |
| rs2236645 | 21 | 33969937 | C | T | 0.17941 | 0.024951 | 6.46E-13 |
| rs2897075 | 7 | 100032719 | C | T | 0.087987 | 0.014375 | 9.32E-10 |
| rs343320 | 3 | 146517122 | G | A | 0.15423 | 0.027508 | 2.06E-08 |
| rs34712979 | 4 | 105897896 | G | A | -0.10951 | 0.01701 | 1.21E-10 |
| rs35705950 | 11 | 1219991 | G | T | -0.1636 | 0.022785 | 6.95E-13 |
| rs368565 | 19 | 48697960 | C | T | 0.1056 | 0.014909 | 1.41E-12 |
| rs41264915 | 1 | 155197995 | A | G | -0.20626 | 0.022896 | 2.09E-19 |
| rs568035 | 1 | 156140376 | C | T | 0.14307 | 0.025967 | 3.60E-08 |
| rs60132559 | 19 | 50363849 | C | T | 0.091456 | 0.015009 | 1.11E-09 |
| rs61882275 | 11 | 34482745 | G | A | -0.12605 | 0.014659 | 8.08E-18 |
| rs62056905 | 17 | 45705327 | A | G | -0.1285 | 0.016821 | 2.18E-14 |
| rs9636867 | 21 | 33237639 | A | G | 0.18394 | 0.015088 | 3.47E-34 |

**Footnote:** IVs - instrumental variables; rsID - variants ID; Chr - Chromosome; Pos - base pair location; EA - effect allele; NEA - non-effect allele; SE - standard error.

**Supplementary Table 4.** Characteristics of genetic variants used as IVs for COVID-19 infection in the East Asian MR analysis

| rsID | Chr | Pos (GRCh38) | EA | NEA | Beta | SE | P-value |
|---|---|---|---|---|---|---|---|
| rs10181451 | 2 | 240232940 | G | T | -0.28658 | 5.87E-02 | 1.04E-06 |
| rs12425722 | 12 | 39942545 | T | C | -0.2981 | 6.09E-02 | 1.00E-06 |
| rs12540473 | 7 | 32523257 | G | A | -0.29579 | 6.35E-02 | 3.20E-06 |
| rs12960174 | 18 | 79832807 | C | T | -0.42397 | 9.17E-02 | 3.75E-06 |
| rs183690023 | 8 | 108100987 | G | A | 0.64177 | 1.32E-01 | 1.08E-06 |
| rs315664 | 4 | 25409021 | T | C | -0.1667 | 3.37E-02 | 7.30E-07 |
| rs76497367 | 2 | 176674648 | C | A | 0.32598 | 7.05E-02 | 3.79E-06 |

**Footnote:** IVs - instrumental variables; rsID - variants ID; Chr - Chromosome; Pos - base pair location; EA - effect allele; NEA - non-effect allele; SE - standard error.

**Supplementary Table 5.** Characteristics of genetic variants used as IVs for hospitalized COVID-19 in the East Asian MR analysis

| rsID | Chr | Pos (GRCh38) | EA | NEA | Beta | SE | P-value |
|---|---|---|---|---|---|---|---|
| rs111553899 | 8 | 42437504 | G | A | -0.42139 | 8.63E-02 | 1.03E-06 |
| rs11919142 | 3 | 192159458 | G | A | 0.36853 | 7.94E-02 | 3.45E-06 |
| rs12175265 | 6 | 41534096 | G | A | 0.273 | 3.76E-02 | 3.73E-13 |
| rs144808322 | 17 | 39274612 | T | G | 0.97979 | 2.10E-01 | 3.15E-06 |
| rs315666 | 4 | 25387180 | G | A | -0.18511 | 3.86E-02 | 1.61E-06 |
| rs4853613 | 2 | 191546927 | T | C | -0.20545 | 4.42E-02 | 3.42E-06 |
| rs72812493 | 5 | 110201691 | A | C | 0.87165 | 1.80E-01 | 1.29E-06 |
| rs73974486 | 2 | 175884081 | A | G | -0.19751 | 4.18E-02 | 2.34E-06 |
| rs77840073 | 10 | 98430961 | C | T | 0.33122 | 7.04E-02 | 2.56E-06 |

**Footnote:** IVs - instrumental variables; rsID - variants ID; Chr - Chromosome; Pos - base pair location; EA - effect allele; NEA - non-effect allele; SE - standard error. One SNP (rs11066150) was excluded following MR-PRESSO outlier detection (Global test P < 0.005)

**Supplementary Table 6.** Characteristics of genetic variants used as IVs for very severe COVID-19 in the East Asian MR analysis

| rsID | Chr | Pos (GRCh38) | EA | NEA | Beta | SE | P-value |
|---|---|---|---|---|---|---|---|
| rs10514025 | 18 | 71827674 | A | C | -0.32208 | 6.78E-02 | 2.06E-06 |
| rs11044248 | 12 | 18687194 | G | A | 0.94413 | 2.05E-01 | 4.08E-06 |
| rs12244041 | 10 | 131476908 | T | C | -0.29985 | 6.34E-02 | 2.26E-06 |
| rs180913314 | 1 | 157288263 | A | G | 0.90471 | 1.86E-01 | 1.17E-06 |
| rs35727104 | 12 | 55110014 | T | C | 0.8991 | 1.91E-01 | 2.39E-06 |
| rs392108 | 6 | 4675766 | C | T | 0.39776 | 7.81E-02 | 3.55E-07 |
| rs55826734 | 9 | 82499301 | A | C | 1.1261 | 2.28E-01 | 7.65E-07 |
| rs75026182 | 9 | 33589677 | C | T | 0.44817 | 9.78E-02 | 4.56E-06 |
| rs76383282 | 14 | 97311620 | G | T | 1.2289 | 2.57E-01 | 1.80E-06 |

**Footnote:** IVs - instrumental variables; rsID - variants ID; Chr - Chromosome; Pos - base pair location; EA - effect allele; NEA - non-effect allele; SE - standard error.

**Supplementary Table 7.** Characteristics of genetic variants used as IVs for COVID-19 infection in the South Asian MR analysis

| rsID | Chr | Pos (GRCh38) | EA | NEA | Beta | SE | P-value |
|---|---|---|---|---|---|---|---|
| rs10880920 | 12 | 46118475 | C | T | -0.08861 | 1.82E-02 | 1.19E-06 |
| rs113893807 | 1 | 47719596 | C | T | -0.24346 | 5.10E-02 | 1.84E-06 |
| rs11629932 | 15 | 69243619 | T | C | -0.0911 | 1.84E-02 | 7.67E-07 |
| rs12523613 | 5 | 37872216 | A | G | 0.097979 | 1.92E-02 | 3.48E-07 |
| rs12669003 | 7 | 120852644 | T | C | 0.58226 | 1.14E-01 | 3.59E-07 |
| rs140619757 | 5 | 12377565 | C | T | 0.39295 | 8.47E-02 | 3.54E-06 |
| rs2122271 | 13 | 57450358 | T | C | -0.12446 | 2.69E-02 | 3.69E-06 |
| rs333098 | 18 | 7176782 | C | T | 0.10745 | 2.28E-02 | 2.54E-06 |
| rs370786079 | 13 | 93939581 | A | C | 0.23853 | 4.90E-02 | 1.12E-06 |
| rs3829653 | 19 | 51713078 | T | G | 0.092025 | 2.00E-02 | 4.40E-06 |
| rs6601370 | 8 | 9827254 | C | T | 0.10366 | 2.10E-02 | 7.66E-07 |
| rs7085134 | 10 | 5549590 | A | G | -0.09998 | 2.07E-02 | 1.44E-06 |
| rs73166412 | 7 | 152615462 | A | G | -0.20117 | 4.37E-02 | 4.23E-06 |
| rs75110192 | 22 | 48873033 | T | G | 0.16291 | 3.52E-02 | 3.76E-06 |
| rs79298572 | 4 | 180565169 | T | C | 0.1609 | 3.50E-02 | 4.38E-06 |

**Footnote:** IVs - instrumental variables; rsID - variants ID; Chr - Chromosome; Pos - base pair location; EA - effect allele; NEA - non-effect allele; SE - standard error.

**Supplementary Table 8.** Characteristics of genetic variants used as IVs for hospitalized COVID-19 in the South Asian MR analysis

| rsID | Chr | Pos (GRCh38) | EA | NEA | Beta | SE | P-value |
|---|---|---|---|---|---|---|---|
| rs1131964 | 21 | 33238182 | T | C | 0.28877 | 4.52E-02 | 1.75E-10 |
| rs12332537 | 5 | 2793392 | A | G | 0.34364 | 7.29E-02 | 2.41E-06 |
| rs1233691 | 6 | 28186119 | T | G | 0.23939 | 4.77E-02 | 5.23E-07 |
| rs12749286 | 1 | 96040592 | G | T | 0.28336 | 6.03E-02 | 2.59E-06 |
| rs12816877 | 12 | 77637000 | T | G | -0.37087 | 7.95E-02 | 3.05E-06 |
| rs299806 | 1 | 79942296 | C | T | -0.4215 | 9.16E-02 | 4.15E-06 |
| rs35032356 | 8 | 94965256 | G | T | 0.47948 | 9.47E-02 | 4.12E-07 |
| rs3742238 | 13 | 112886111 | C | T | 0.246 | 5.00E-02 | 8.48E-07 |
| rs374304656 | 4 | 24912655 | G | A | 1.112 | 2.42E-01 | 4.50E-06 |
| rs571136 | 6 | 108757208 | C | T | -0.22192 | 4.71E-02 | 2.50E-06 |
| rs57885096 | 10 | 86144539 | C | T | 0.26635 | 5.55E-02 | 1.62E-06 |
| rs6660884 | 1 | 155053638 | C | T | -0.27799 | 4.51E-02 | 6.80E-10 |
| rs72787814 | 16 | 63048846 | G | A | -0.51941 | 1.13E-01 | 4.03E-06 |
| rs74401283 | 6 | 105520426 | T | C | 0.94515 | 2.05E-01 | 4.12E-06 |
| rs75968133 | 4 | 54703444 | C | T | 0.90476 | 1.87E-01 | 1.26E-06 |
| rs76681218 | 3 | 127652778 | T | C | 0.94014 | 2.01E-01 | 2.85E-06 |
| rs80175346 | 16 | 89191507 | T | C | 0.48193 | 9.79E-02 | 8.46E-07 |
| rs8068105 | 17 | 76662002 | G | T | 0.30044 | 6.41E-02 | 2.81E-06 |

**Footnote:** IVs - instrumental variables; rsID - variants ID; Chr - Chromosome; Pos - base pair location; EA - effect allele; NEA - non-effect allele; SE - standard error.

**Supplementary Table 9.** Characteristics of genetic variants used as IVs for very severe COVID-19 in the South Asian MR analysis

| rsID | Chr | Pos (GRCh38) | EA | NEA | Beta | SE | P-value |
|---|---|---|---|---|---|---|---|
| rs1131964 | 21 | 33238182 | T | C | 0.26943 | 5.41E-02 | 6.30E-07 |
| rs116734871 | 1 | 237943396 | T | C | 0.57475 | 1.25E-01 | 4.39E-06 |
| rs12332537 | 5 | 2793392 | A | G | 0.41143 | 8.53E-02 | 1.42E-06 |
| rs1233691 | 6 | 28186119 | T | G | 0.30319 | 5.71E-02 | 1.08E-07 |
| rs13078854 | 3 | 45820440 | G | A | 0.69791 | 6.11E-02 | 3.01E-30 |
| rs13118628 | 4 | 169973300 | T | C | -0.24563 | 5.36E-02 | 4.52E-06 |
| rs138246166 | 7 | 3184731 | A | C | 1.2681 | 2.63E-01 | 1.41E-06 |
| rs141138593 | 10 | 59208951 | T | C | 0.58141 | 1.26E-01 | 4.09E-06 |
| rs147653711 | 16 | 89206191 | C | T | 0.57577 | 1.06E-01 | 5.69E-08 |
| rs191630535 | 20 | 49201977 | T | C | 0.54703 | 1.15E-01 | 1.84E-06 |
| rs571136 | 6 | 108757208 | C | T | -0.25586 | 5.57E-02 | 4.30E-06 |
| rs578709 | 6 | 78689867 | T | C | 0.25255 | 5.44E-02 | 3.41E-06 |
| rs59395680 | 12 | 58405677 | T | C | -0.47997 | 1.05E-01 | 4.58E-06 |
| rs6660884 | 1 | 155053638 | C | T | -0.28896 | 5.41E-02 | 9.22E-08 |
| rs73958215 | 18 | 31842164 | A | G | 0.88974 | 1.76E-01 | 4.00E-07 |
| rs75968133 | 4 | 54703444 | C | T | 1.0513 | 2.12E-01 | 7.29E-07 |
| rs76150991 | 12 | 100460195 | A | C | 0.35141 | 7.54E-02 | 3.16E-06 |
| rs76773794 | 21 | 45227898 | A | G | -0.67614 | 1.45E-01 | 3.24E-06 |
| rs8134143 | 21 | 20609358 | G | A | -0.46198 | 9.63E-02 | 1.59E-06 |

**Footnote:** IVs - instrumental variables; rsID - variants ID; Chr - Chromosome; Pos - base pair location; EA - effect allele; NEA - non-effect allele; SE - standard error.

**Supplementary Table 10.** Characteristics of genetic variants used as IVs for COVID-19 infection in the African MR analysis

| rsID | Chr | Pos (GRCh38) | EA | NEA | Beta | SE | P-value |
|---|---|---|---|---|---|---|---|
| rs112050567 | 16 | 29162237 | C | T | -0.23715 | 4.99E-02 | 2.05E-06 |
| rs114377730 | 5 | 168768842 | C | T | -0.42299 | 9.15E-02 | 3.79E-06 |
| rs114623037 | 4 | 2319211 | G | A | 0.43366 | 8.83E-02 | 8.93E-07 |
| rs116773716 | 20 | 54075638 | C | T | 0.39066 | 8.44E-02 | 3.70E-06 |
| rs138712344 | 10 | 98868625 | C | T | 0.45201 | 9.66E-02 | 2.91E-06 |
| rs138865339 | 7 | 157682288 | G | A | 0.23019 | 4.91E-02 | 2.76E-06 |
| rs139701757 | 17 | 54031675 | T | C | 0.43149 | 9.33E-02 | 3.75E-06 |
| rs1447202 | 2 | 142544380 | C | T | 0.090271 | 1.86E-02 | 1.16E-06 |
| rs146539894 | 9 | 87189786 | G | A | 0.3654 | 7.46E-02 | 9.52E-07 |
| rs148020121 | 10 | 92387831 | A | G | -0.28737 | 5.95E-02 | 1.36E-06 |
| rs16888850 | 4 | 13578987 | T | C | 0.10882 | 2.33E-02 | 2.95E-06 |
| rs17565505 | 5 | 152684873 | C | T | 0.1784 | 3.63E-02 | 9.15E-07 |
| rs186054942 | 3 | 73347665 | G | A | 0.58193 | 1.21E-01 | 1.57E-06 |
| rs2268147 | 1 | 201283738 | C | T | -0.0835 | 1.77E-02 | 2.36E-06 |
| rs2571228 | 18 | 57773827 | C | A | -0.2533 | 5.43E-02 | 3.08E-06 |
| rs2823960 | 21 | 16673220 | T | C | 0.08164 | 1.75E-02 | 2.91E-06 |
| rs376454629 | 6 | 141497237 | G | A | 0.41223 | 8.52E-02 | 1.30E-06 |
| rs393507 | 5 | 101468720 | C | T | 0.096716 | 1.67E-02 | 7.34E-09 |
| rs4623022 | 4 | 125412242 | C | T | 0.094422 | 1.91E-02 | 8.10E-07 |
| rs571364782 | 14 | 76331899 | T | C | 0.44619 | 9.25E-02 | 1.39E-06 |
| rs62574963 | 9 | 108642439 | A | G | -0.15482 | 3.38E-02 | 4.58E-06 |
| rs6496411 | 15 | 87338824 | G | A | 0.17103 | 3.48E-02 | 8.87E-07 |
| rs73276601 | 14 | 63505215 | C | A | -0.44702 | 9.13E-02 | 9.66E-07 |
| rs73592155 | 13 | 104006656 | A | C | 0.22707 | 4.97E-02 | 4.86E-06 |
| rs743567 | 14 | 23421773 | T | G | -0.08545 | 1.82E-02 | 2.55E-06 |
| rs75992633 | 16 | 71872721 | G | T | 0.42084 | 8.77E-02 | 1.59E-06 |
| rs79039808 | 4 | 67938190 | G | T | 0.20882 | 4.40E-02 | 2.10E-06 |

**Footnote:** IVs - instrumental variables; rsID - variants ID; Chr - Chromosome; Pos - base pair location; EA - effect allele; NEA - non-effect allele; SE - standard error.

**Supplementary Table 11.** Characteristics of genetic variants used as IVs for hospitalized COVID-19 in the African MR analysis

| rsID | Chr | Pos (GRCh38) | EA | NEA | Beta | SE | P-value |
|---|---|---|---|---|---|---|---|
| rs10023625 | 4 | 187891010 | C | A | 0.15532 | 3.14E-02 | 7.50E-07 |
| rs10402036 | 19 | 32988607 | T | C | -0.36788 | 8.00E-02 | 4.25E-06 |
| rs114007484 | 4 | 61110486 | T | C | -0.67384 | 1.39E-01 | 1.25E-06 |
| rs114442516 | 12 | 46143049 | T | C | 0.33227 | 7.09E-02 | 2.75E-06 |
| rs115033748 | 1 | 155783076 | A | G | -0.33816 | 6.86E-02 | 8.35E-07 |
| rs116437405 | 5 | 109503100 | T | G | 0.65744 | 1.40E-01 | 2.57E-06 |
| rs116538606 | 14 | 44076407 | T | C | 0.68843 | 1.49E-01 | 3.87E-06 |
| rs11873691 | 18 | 65153912 | C | T | 0.64646 | 1.22E-01 | 1.08E-07 |
| rs12831587 | 12 | 18223818 | T | C | 0.41661 | 8.93E-02 | 3.05E-06 |
| rs138445399 | 4 | 78722572 | A | C | 0.65029 | 1.40E-01 | 3.54E-06 |
| rs140546786 | 14 | 54672243 | T | C | 0.40717 | 8.11E-02 | 5.11E-07 |
| rs144144723 | 4 | 14950134 | C | T | 0.67015 | 1.37E-01 | 9.30E-07 |
| rs145249745 | 11 | 114491290 | C | T | 0.36567 | 7.77E-02 | 2.56E-06 |
| rs146891917 | 8 | 100506982 | C | T | 0.61759 | 1.32E-01 | 2.84E-06 |
| rs147219386 | 3 | 170652421 | A | G | 0.60649 | 1.23E-01 | 8.56E-07 |
| rs148255307 | 4 | 85782921 | G | A | 0.50613 | 1.08E-01 | 2.55E-06 |
| rs190134128 | 1 | 25809896 | G | A | 0.33938 | 6.38E-02 | 1.02E-07 |
| rs2331294 | 22 | 26699747 | G | A | 0.14778 | 3.13E-02 | 2.35E-06 |
| rs34487659 | 8 | 40061538 | G | A | -0.28821 | 6.24E-02 | 3.93E-06 |
| rs56095707 | 21 | 45464981 | A | C | 0.63138 | 1.35E-01 | 2.76E-06 |
| rs61687419 | 3 | 136112014 | G | A | 0.24604 | 5.10E-02 | 1.38E-06 |
| rs6486606 | 12 | 130852781 | T | G | -0.19564 | 4.21E-02 | 3.30E-06 |
| rs7080718 | 10 | 125069355 | C | T | 0.18098 | 3.77E-02 | 1.61E-06 |
| rs74393060 | 2 | 162987285 | A | C | 0.24712 | 5.40E-02 | 4.74E-06 |
| rs75268797 | 5 | 3029047 | A | G | 0.78966 | 1.42E-01 | 2.63E-08 |
| rs78605069 | 20 | 17256495 | T | G | 0.29357 | 6.03E-02 | 1.12E-06 |
| rs825267 | 2 | 221701197 | C | A | 0.16041 | 3.51E-02 | 4.90E-06 |

**Footnote:** IVs - instrumental variables; rsID - variants ID; Chr - Chromosome; Pos - base pair location; EA - effect allele; NEA - non-effect allele; SE - standard error.

**Supplementary Table 12.** Characteristics of genetic variants used as IVs for very severe COVID-19 in the African MR analysis

| rsID | Chr | Pos (GRCh38) | EA | NEA | Beta | SE | P-value |
|---|---|---|---|---|---|---|---|
| rs10176143 | 2 | 1E+08 | A | G | -0.31515 | 0.066972 | 2.53E-06 |
| rs115408289 | 4 | 79588758 | T | C | 0.82274 | 0.17515 | 2.63E-06 |
| rs116771446 | 4 | 1.77E+08 | A | G | -0.96478 | 0.20524 | 2.59E-06 |
| rs13151613 | 4 | 48255598 | G | T | 0.40693 | 0.088365 | 4.12E-06 |
| rs13223986 | 7 | 14463928 | G | A | -0.34052 | 0.073582 | 3.70E-06 |
| rs138194738 | 6 | 1.62E+08 | T | C | 1.2291 | 0.25562 | 1.52E-06 |
| rs146243869 | 12 | 20660726 | C | T | 0.85288 | 0.18649 | 4.80E-06 |
| rs17038551 | 2 | 49264427 | A | G | -0.33229 | 0.070415 | 2.37E-06 |
| rs67295842 | 18 | 40569023 | C | T | 0.57476 | 0.12522 | 4.43E-06 |
| rs9586580 | 13 | 1.05E+08 | G | A | 0.92937 | 0.20236 | 4.37E-06 |

**Footnote:** IVs - instrumental variables; rsID - variants ID; Chr - Chromosome; Pos - base pair location; EA - effect allele; NEA - non-effect allele; SE - standard error.
